# Real-World Improvement in Motor Status Among Regular Users of a Home-Based Brain-Computer Interface After Stroke

**DOI:** 10.64898/2026.10.30.26366945

**Authors:** Phillip Demarest, Steven C. Cramer, Gregory E. Wilding, Nancey Trevanian Tsai, Sarah Gibbons, Nikhita J. Perry, Lee A. Fleisher, Eric C. Leuthardt, Seth J. Wilk

## Abstract

**Background:** Upper extremity impairment persists in many stroke survivors, and few treatments are effective for severe chronic deficits. Brain-computer interface (BCI) therapy, which pairs decoded motor-intention-associated electrophysiology with sensory feedback, has improved upper extremity impairment in randomized trials, including in patients with severe paresis. Whether these benefits translate to routine care has rarely been evaluated. This study examined upper extremity motor recovery through 1 year among patients prescribed at-home BCI therapy in clinical practice.

**Methods:** This retrospective observational study included adults who were prescribed the IpsiHand System, an electroencephalography-based BCI, through a commercial US home therapy program, had a baseline Upper Extremity Fugl-Meyer (UEFM) assessment, and averaged >1 session per week through their last assessment. Occupational therapists administered the UEFM by video every 4 weeks. The primary outcome was UEFM change from baseline to week 12. A mixed model for repeated measures estimated change through week 52, with multiple imputation sensitivity analyses for missing data.

**Results:** Of 452 patients included in analyses (mean time since stroke, 5.4±5.4 years), mean baseline UEFM was 20.1±14.6, with 333 patients (73.7%) having severe motor impairment (UEFM 0-28). At week 12, mean UEFM change was 4.3 points (95% CI, 3.7–4.9; P<0.001), and 31.7% achieved the minimal clinically important difference (MCID) of 5.25 points. Mean change reached the MCID at week 16 (5.6, 95% CI [4.8, 6.4]). Model-estimated change in UEFM continued to improve through 1 year, being 6.1 (95% CI, 5.5–6.8) at week 24 and 8.1 (95% CI, 6.8–9.4) at week 52.

**Conclusions:** Among regular users of at-home BCI therapy in routine care, most of whom had severe chronic impairment, UEFM scores improved by 12 weeks, reached MCID by 16 weeks, and continued to increase through 1 year. At-home BCI therapy merits evaluation in randomized trials with follow-up beyond 12 weeks.

## Introduction

Stroke is highly debilitating and a leading cause of death, with an estimated 795,000 cases in the United States annually and an estimated 12 million globally each year.^1,2^ Upper-limb impairment persists in about 40% of survivors, significantly impacting their daily living and quality of life.^3,4^ Current treatment modalities for functional rehabilitation in chronic stroke survivors are limited. Few stroke survivors receive physical or occupational therapy in the chronic phase,^5^ and while impairment can still be reduced with rehabilitation,^6–8^ this may require therapy doses that are more intensive and not often feasible.^9–11^ Further, accessibility of therapies can be difficult, particularly for patients who require at-home solutions.^12–15^

Brain-computer interface (BCI) technologies provide a promising solution for chronic stroke rehabilitation.^16,17^ BCI systems can be noninvasive, and in some cases use portable hardware elements that can be safely deployed for at-home use.^18–21^ Therapeutic BCIs operate by decoding target electrophysiological biomarkers and concurrently driving physiologically relevant neurofeedback to promote beneficial neural remodeling to promote recovery.^22^ This form of BCI-mediated cortico-sensory coupling is thought to induce Hebbian learning by simultaneously activating pre-and post-synaptic neurons. In the context of stroke, concurrent reinforcement of physiological mechanisms related to motor intention through assisted motor feedback has been shown to promote motor function and lead to long-term changes in neurophysiology.^20,23^

Prior studies applying BCI systems for chronic stroke recovery have reported motor recovery benefits and have demonstrated reliable BCI control in patients with hemiparesis through decoding of motor-related electroencephalographic (EEG) signals from the contralesional hemisphere, ipsilateral to the paretic limb.^20^ In a pooled analysis of two prospective single-arm studies, 12 weeks of BCI therapy increased patient Upper Extremity Fugl-Meyer (UEFM) scores by a mean of 8.1 points.^24^ In the recent BCI-REHAB randomized clinical trial (Unique identifier: NCT05965713), patients receiving BCI therapy averaged a 6.0 UEFM improvement after 12 weeks compared to a 1.5 UEFM improvement in the home-exercise control group, with over half of participants assigned to BCI therapy achieving clinically meaningful outcomes.^25^ The outcomes from these studies not only support the therapeutic efficacy of BCI systems for motor recovery but also demonstrate the feasibility and accessibility of using such systems in a home environment. Whether these gains are enduring, and whether benefits translate to non-clinical research settings, however, remains uncertain. These studies assessed primary outcomes at the end of 12 weeks of therapy. Across BCI therapies for stroke, a meta-analysis found immediate benefits,^26^ but only two trials reported gains that were maintained 6 to 12 months after therapy ended.^27,28^

While such studies have provided promising evidence for the benefits of at-home BCI systems for chronic stroke motor rehabilitation, they were conducted using prospective protocols, and real-world evidence derived from data collected during standard clinical care remains limited.^29^ Understanding whether the benefits observed in prospective trials carry over to routine clinical care is critical for guiding appropriate adoption of these systems of care and for setting realistic expectations of benefit for patients and providers.^30,31^ The present study provides a single-arm, observational characterization of upper extremity motor impairment changes in a population of chronic stroke survivors utilizing an at-home BCI therapy system embedded within routine care. The goal of this study is to provide real-world evidence of upper extremity motor recovery with at-home BCI therapy by (1) quantifying UEFM changes after 12 weeks of use and (2) characterizing the trajectory of change through one year.

## Methods

### Study design, ethics, and data governance

This retrospective study presents a single-arm, observational characterization of the changes in UEFM within a population of real-world BCI system users. Patients were prescribed the IpsiHand BCI system in accordance with its FDA-cleared indication (FDA De Novo DEN200046; 21 CFR 890.5420) as part of routine care. Data were extracted from clinical and administrative records and from device session logs. Direct identifiers were removed before any analysis. The dataset was frozen on September 6, 2026; the UEFM assessments analyzed were recorded between May 2023 and September 6, 2026. Social Deprivation Index (SDI) was determined using patient ZIP code information.^32^ Time since stroke was calculated from the recorded stroke date to the baseline assessment date. This study was deemed exempt from ethics review by the Western Institutional Review Board-Copernicus Group (tracking number 20250386) on February 7, 2025, under US federal regulations (45 CFR 46.104(d)(4)) as secondary research using existing records. All patients provided consent, although the exemption did not require it. Study design and the statistical methods used in this study were defined prior to analysis in a statistical analysis plan, which can be found in Supplemental Materials. This study is reported according to the STROBE guideline and the checklist can be found in Supplemental Materials.

### Patient population

This study includes data from patients who (1) were 18 years and older and were prescribed the IpsiHand between January 1 2023 and September 6, 2026, (2) had a stroke at least six months before beginning IpsiHand therapy, and (3) had hemiparesis or hemiplegia. Exclusion criteria were (1) absence of a baseline UEFM measurement, (2) failure to use the device for an average of more than one session per week averaged over all weeks between baseline and each respective patient’s most recent follow-up assessment date, (3) any significant detrimental medical event after starting IpsiHand therapy, defined as an event, injury, or change in neurological or functional status that may compromise the patient’s safety or ability to continue therapy, including but not limited to recurrent stroke, hospitalization, fall with injury, fracture, or surgery, and (4) impaired cognitive function, language, motor function, or visual attention indicative of an inability to use the device properly or follow instructions. The device-use criterion was applied so that outcomes described patients who used the device regularly. Every logged session with a duration greater than zero counted toward the average, and for patients without a follow-up assessment, the average extended to the data cutoff. No sample size was calculated, and all eligible patients were included.

Before initiating therapy, candidates completed an EEG signal test confirming that the system could detect a control signal over the contralesional hemisphere. Program clinical staff scheduled the test, which was performed either in a clinic or in the patient’s home. The signals were evaluated as described previously.^20,24,25^ In sum, a dry-electrode EEG headset was placed with electrodes at frontal, central, and parietal sites of the international 10-20 system, including C3 and C4 over the left and right sensorimotor cortex. Patients performed a visually cued task of rest and motor imagery trials and were instructed to imagine moving the affected hand.

Spectral power was estimated with an autoregressive method, and at each electrode and frequency, the coefficient of determination (r^2^) quantified the proportion of variance in spectral power explained by the difference between imagery and rest trials. Specifically, this was the squared correlation between power and trial type. A usable control signal was defined as an event-related desynchronization in the mu (8-12 Hz) or beta (13-29 Hz) band at an electrode over the contralesional sensorimotor cortex, consistent across repeated recordings. The electrode and the frequency bin (width 1 Hz) with the strongest r^2^ became the patient’s control feature and remained fixed for the course of therapy. Candidates without a usable control signal did not proceed to therapy.

### Intervention, setting, and care pathway

IpsiHand is a non-invasive BCI therapy for patients undergoing stroke rehabilitation to facilitate and to maintain or increase upper-extremity range of motion.^33^ The system consists of a wireless EEG headset that records neural activity over both sensorimotor cortices, a tablet that runs the therapy software, and a motorized exoskeleton worn over the affected hand and wrist. Signal recordings are decoded in real time by estimating spectral power at the patient’s control feature determined during EEG screening. When spectral features of imagined movement (event-related desynchronization in 8-12 Hz and 12-30 Hz power) are detected, a control signal is transmitted to the motorized exoskeleton, which opens and closes the hand in a three-finger pinch and which also provides proprioceptive feedback. A flow diagram of the BCI system is shown in Figure 1A.

**Figure 1.**
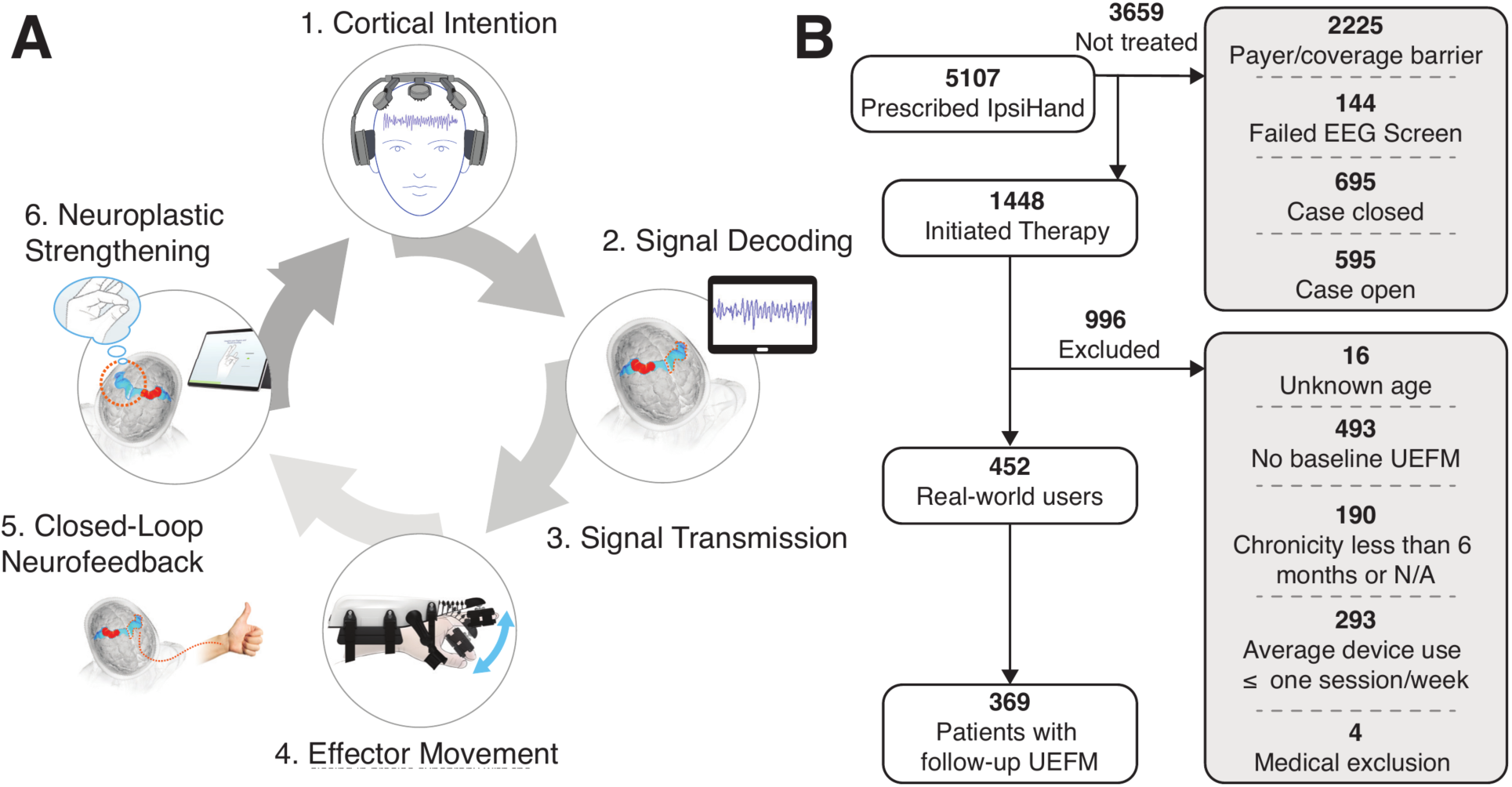
Overview of the brain-computer interface (BCI) system and the real-world subpopulation. (**A**) Flow diagram of the BCI system. The patient initiates motor intention to move the affected limb. Corresponding electrophysiological activity is detected over the motor cortex with a wireless EEG headset. The decoded motor signals are transmitted as control signals to the handpiece effector, which opens and closes the fingers of the affected extremity in a 3-finger pinch concurrently with motor intention. This BCI-mediated closed-loop neurofeedback training is proposed to support therapeutic neural remodeling with repeated use. (**B**) Profile of patients included in and excluded from the present study.

Each BCI therapy session consisted of a calibration phase followed by five blocks of 60 trials, with a full session lasting roughly 1 hour. During calibration, patients first remained at rest for 3 minutes, then completed a set of motor imagery and rest trials while the exoskeleton remained stationary. Calibration was used to ensure data quality and to identify motor imagery activity. Each block comprised 30 motor imagery trials and 30 rest trials in a randomized order. Trials lasted 8 seconds and were separated by 3-second buffer intervals during which patients were encouraged to blink or make minor physical adjustments. Blocks were separated by pauses during which patients could rest before resuming therapy. During motor imagery trials, patients were instructed to imagine opening the affected hand. In rest trials, patients were instructed to keep the exoskeleton closed by resting quietly.

Patients received the device through a clinical program administered by Kandu, Inc. (Van Nuys, Cam USA). Each patient first attended a consultation with the program’s clinical team to confirm eligibility, which required a stroke at least six months prior to consultation, age of 18 years or older, persistent upper-extremity motor impairment, and the ability to participate in a home-based program. Patients who met these criteria then obtained a physician prescription. The prescription and supporting medical records were submitted for payer authorization, or patients purchased the device through self-pay. Before initiating therapy, patients were also required to pass a set of screening procedures to ensure they met device eligibility, which included a cognitive screen, a clinical upper-extremity assessment, and the EEG signal test described in *Patient population*. Patients without aphasia completed the Short Blessed Test,^34^ and patients with aphasia were assessed with the Mississippi Aphasia Screening Test.^35^ All patients were screened for visual inattention with an unstructured cancellation test.^36^ Patients initiating therapy received training and guidance on how to don and use the device from a licensed occupational therapist. Therapy was self-administered at home, and patients chose when and how often to use the device. Patients were provided device technical support through the clinical program by video conference or video call.

### Outcome and assessment

The primary outcome was the UEFM total score (range 0 to 66, where higher scores indicate less impairment) during follow-up assessments up to 52 weeks. Week 12 was specified as the primary outcome timepoint because it corresponded to the treatment duration and endpoint of interest in published prospective studies of this device.^20,24,25^ Assessments were administered remotely using videoconference, by licensed occupational therapists trained and certified in the Fugl-Meyer assessment.^37,38^ Assessors knew patients were using the device and were therefore unblinded. Training and certification included completion of the standardized BlueCloud Fugl-Meyer training, followed by at least three remotely administered Fugl-Meyer assessments reviewed and independently scored by a mentor clinician.

Initial baseline UEFM assessments were conducted during onboarding and training after patients received the device. Follow-up assessments were conducted on a 4-week schedule; however, variation in assessment dates occurred due to patient availability. For analysis, follow-up assessments were organized into 4-week bins (week ± 2 weeks).

UEFM change was calculated as the difference between the follow-up score and the baseline score. A ≥ 5.25 change in UEFM was chosen as the minimal clinically important difference (MCID) threshold for this study based on existing literature.^39^ Severity categories were determined as UEFM total score 0-28 for severe, 29-42 for moderate, and 43-66 for mild.^40^

### Statistical analysis

Analyses were performed in Python 3.9.6 (pandas 2.3.3, NumPy 2.0.2, and SciPy 1.13.1) and R 4.6.1 (mmrm 0.3.18). Change from baseline at each assessment week was summarized among patients with both a baseline and respective follow-up measure by the mean and its 95% confidence interval (CI) based on the t distribution, a 95% patient-level bootstrap CI, the median and the interquartile range, Cohen’s d_z_ as a standardized measure of effect size appropriate for paired data, percentage of patients showing improvement from baseline (UEFM change > 0), and the percentage of subjects with UEFM change that reached or exceeded the MCID. A paired t-test was used to evaluate significance of UEFM differences between baseline and follow-up assessment weeks. Paired t-test p values were adjusted across all paired comparisons (n = 13) by the Benjamini-Hochberg (BH) procedure. An adjusted p < 0.05 was considered significant.

Overall UEFM trajectory across available follow-up assessments up to a year was modeled using a mixed model for repeated measures (MMRM) to characterize changes in UEFM after the primary follow-up date. This model included every patient with at least one follow-up assessment, and UEFM at each assessment week was the response. The model had two fixed effects: assessment week, entered as an unordered categorical variable, and baseline UEFM. Baseline UEFM was included to account for differences in baseline impairment among the patients assessed at each week. No random effects were included. Correlation between each patient’s repeated assessments was modeled through the covariance of the within-patient residuals. The model was fitted by restricted maximum likelihood, and inference on the fixed effects used Kenward-Roger degrees of freedom.^41^ Due to variance in follow-up trajectories, and patient factors, data were assumed missing at random and missing values were handled by direct likelihood.^42^

For each patient *i* at follow-up visit *t*, the UEFM total score was modeled as

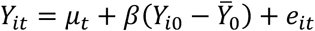

where *Y_it_*is patient *i*‘s UEFM score at observation *t*, *μ_t_* is the least-squares mean UEFM at week *t*, adjusted to the mean baseline, *Y*_*i*0_ is the patient’s baseline score, *Y̅*_0_ is the mean baseline UEFM from the analysis set, *β* is the regression slope on baseline UEFM, and *e_it_* is the within-patient residual. Each patient’s residuals were assumed to be multivariate normal with mean zero and a heterogeneous first-order ante-dependence covariance matrix, which has a separate variance for each of the 13 weeks and a separate correlation for each pair of consecutive weeks.

Two additional sensitivity analyses were conducted to examine the missing-at-random assumption of the MMRM. Each analysis involved imputation of due but absent assessments 50 times and pooling the results by Rubin’s rules.^43^ First, a carry-forward multiple imputation replaced each due but absent assessment with the patient’s last observed value plus a zero-mean normal draw with the model’s week-specific standard deviation. The MMRM was then refitted to each completed dataset. Second, a tipping-point analysis was used to impute each assessment under missing at random, from the model’s conditional distribution given the patient’s observed scores. A fixed penalty term (δ) was subtracted from the imputed values after week 12 before refitting. This was repeated over a range of δ to determine the smallest δ at which the 95% confidence interval of the modeled difference between week 12 and week 52 contained 0. This analysis allowed us to determine the magnitude of departure from missing at random required before the improvement from week 12 to week 52 was no longer supported.

### Exploratory analyses

After the primary analyses, we conducted three post hoc exploratory analyses of baseline characteristics. First, we compared age, baseline UEFM, time since stroke, and social deprivation index between patients that improved by at least MCID and those who did not at weeks 12, 16, and 24. Second, we compared baseline characteristics between patients with a week 12 assessment and patients whose 12-week assessment was due but not recorded.

Third, we compared baseline characteristics between patients with at least one follow-up assessment and patients with only a baseline assessment. Statistical comparisons were made using a Wilcoxon rank-sum test, and p-values were adjusted using the BH procedure.

Given the primary results and the predominance of severe baseline impairment in our cohort, we conducted an additional post hoc exploratory analysis of patients with a baseline UEFM of 20-50, the inclusion range of previously reported clinical trials of upper extremity motor recovery after stroke.^44–46^ This range excludes deficits mild enough to produce a ceiling effect and deficits too severe for meaningful recovery to be likely.^47–49^ In these patients, we summarized UEFM change from baseline at weeks 12, 16, and 24 with the statistics used in the primary analysis and adjusted t-test p values across the three weeks using the BH procedure.

### AI use

Anthropic’s Claude and Claude Code, including custom agentic workflows and agent skills were used in preparation of the manuscript and analyses. Agentic workflows were applied to organize citations, make suggestions for included literature, assist with cross-referencing results from existing literature, prepare the STROBE checklist, proofread the final manuscript, and format documents for journal-specific requirements. Agentic workflows were also applied to test, debug, and organize the analysis code, and package the analysis for GitHub.

### Data availability

All code used in this study can be found on GitHub [https://github.com/KanduHealth/ipsihand-rwe-uefm]. The data analyzed in this study are not publicly available to protect patient information. Data may be made available from the corresponding author on reasonable request.

## Results

### Patient characteristics

Out of 5107 patients who received an IpsiHand prescription, 1448 received a device and were cleared to initiated therapy. Of the 3659 patients prescribed IpsiHand, but did not initiate therapy, 2225 were denied payer coverage or faced a payment barrier, 144 failed EEG screening, 695 had their inquiry case closed due to withdrawal or other reasons (including lack of medical records, patient withdrawal, or clinical ineligibility), and 595 were awaiting next steps towards receiving the IpsiHand system. Of these, 16 lacked a documented age or were younger than 18 years, 493 did not have a baseline UEFM assessment, 190 had a stroke chronicity of less than six months or did not have a stroke date documented, 293 averaged one session per week or fewer (mean 0.25, SD 0.28, range [0.00, 1.00]), and 4 had a significant detrimental medical event.

Overall, the full patient cohort included in the analysis set consisted of 452 real-world device users who met our inclusion criteria and received baseline UEFM assessments (mean age 61.9, SD 13.5; 253 male, 185 female of 438 with recorded sex), with 369 of them receiving at least one follow-up assessment (mean age 61.6 SD 13.7; 201 male, 154 female of 355 with recorded sex). In these 369 patients, median follow-up from baseline to the last UEFM assessment was 16.6 weeks (IQ range [9.0, 28.1]). Figure 1B shows the profile of patients included and excluded from the present study. Supplemental Table 1 provides the number of patients with a UEFM assessment at each follow-up week. Supplemental Table 2 provides a breakdown of payer information across patients prescribed the IpsiHand, patients that initiated therapy, and patients included in analysis.

Table 1 summarizes patient characteristics. Most patients had severe upper extremity motor impairment, with mean baseline UEFM 20.1 (SD 14.6), 333 patients (73.7%) having severe motor impairment (UEFM 0-28), 74 (16.4%) with moderate impairment (UEFM 29-42), and 45 (10.0%) with mild impairment (43-66). Mean time since stroke at baseline was 5.4 years (SD 5.4, range [0.50, 39.49]). Overall, patients completed an average of 3.6 sessions per week (SD 1.8, range [1.04 – 10.76]). Notably, while this study evaluates changes in UEFM based on available follow-up assessments, the absence of an outcome assessment does not necessarily mean patients were not continuing to use the device. A table outlining how many patients in the analysis set were using the device during follow-up assessment windows is shown in Supplemental Table 3. Notably, 85.8% of patients who still owned the device were using the device at week 12 (308/359), 77.7% at week 16 (258/332), 71.4% at week 24 (192/269), and 41.3% at 1 year (31/75). However, among the 369 patients with a follow-up assessment, a UEFM assessment was recorded for 52.9% of those due at week 12, 45.8% at week 16, 32.9% at week 24, and 15.8% at 1 year (Supplemental Table 1).

**Table 1.** Patient demographics and baseline characteristics.

|  | **Full patient cohort (N=452)** | **Patients with UEFM follow up<br>(N=369)** |
|---|---|---|
| **Age at baseline, years (mean, SD)** | **61.9 (13.5)** | **61.6 (13.7)** |
| **Sex, n (%)** |  |  |
| Male | 253 (56.0) | 201 (54.5) |
| Female | 185 (40.9) | 154 (41.7) |
| N/A | 14 (3.1) | 14 (3.8) |
| **Baseline UEFM, 0–66 (mean, SD)** | **20.1 (14.6)** | **20.0 (14.4)** |
| **Baseline severity band, n (%)** |  |  |
| Severe (0–28) | 333 (73.7) | 270 (73.2) |
| Moderate (29–42) | 74 (16.4) | 65 (17.6) |
| Mild (43–66) | 45 (10.0) | 34 (9.2) |
| **Time since stroke at baseline, years (mean, SD)** | **5.4 (5.4); n = 451** | **5.3 (5.3)** |
| **Affected upper extremity, n (%)** |  |  |
| Left | 256 (56.6) | 206 (55.8) |
| Right | 196 (43.4) | 163 (44.2) |
| **Dominant hand, n (%)** |  |  |
| Right | 341 (75.4) | 271 (73.4) |
| Left | 43 (9.5) | 38 (10.3) |
| Ambidextrous | 3 (0.7) | 3 (0.8) |
| N/A | 65 (14.4) | 57 (15.4) |
| **Social Deprivation Index, 1–100 (mean, SD)** | **44.5 (27.3); n = 451** | **44.5 (26.9); n = 368** |

### UEFM improvements after 12 weeks

Among the 180 patients with baseline and week-12 assessments, we observed a significant paired mean UEFM increase of 4.3 points (95% CI [3.7, 4.9]; bootstrap 95% CI [3.8, 5.0]; SD 4.1; median 4; IQ range [1, 7], d_z_ 1.06; paired t-test p < 0.001; Figure 2A). Scores improved in 151 patients (83.9%), did not improve in 29 (16.1%), and improved by at least MCID in 57 (31.7%) (Figure 2B). A comparison of baseline characteristics between patients that improved by at least MCID and patients who did not appears in Supplemental Figure 1. Patients who improved by at least the MCID were significantly less impaired at baseline (median baseline UEFM 28.0, IQ range [13.0, 39.0] vs 11.0 [7.0, 24.5]; Wilcoxon rank-sum test BH adjusted p < 0.001). There were no significant differences in age, time since stroke, or SDI. These results were consistent at weeks 16 (median 18.0, IQ range [11.0, 33.0] vs 10.0 [7.0, 20.0]; BH adjusted p < 0.001) and 24 (median 24.5, IQ range [11.0, 33.0] vs 11.0 [7.3, 19.0] BH adjusted p = 0.007).

**Figure 2.**
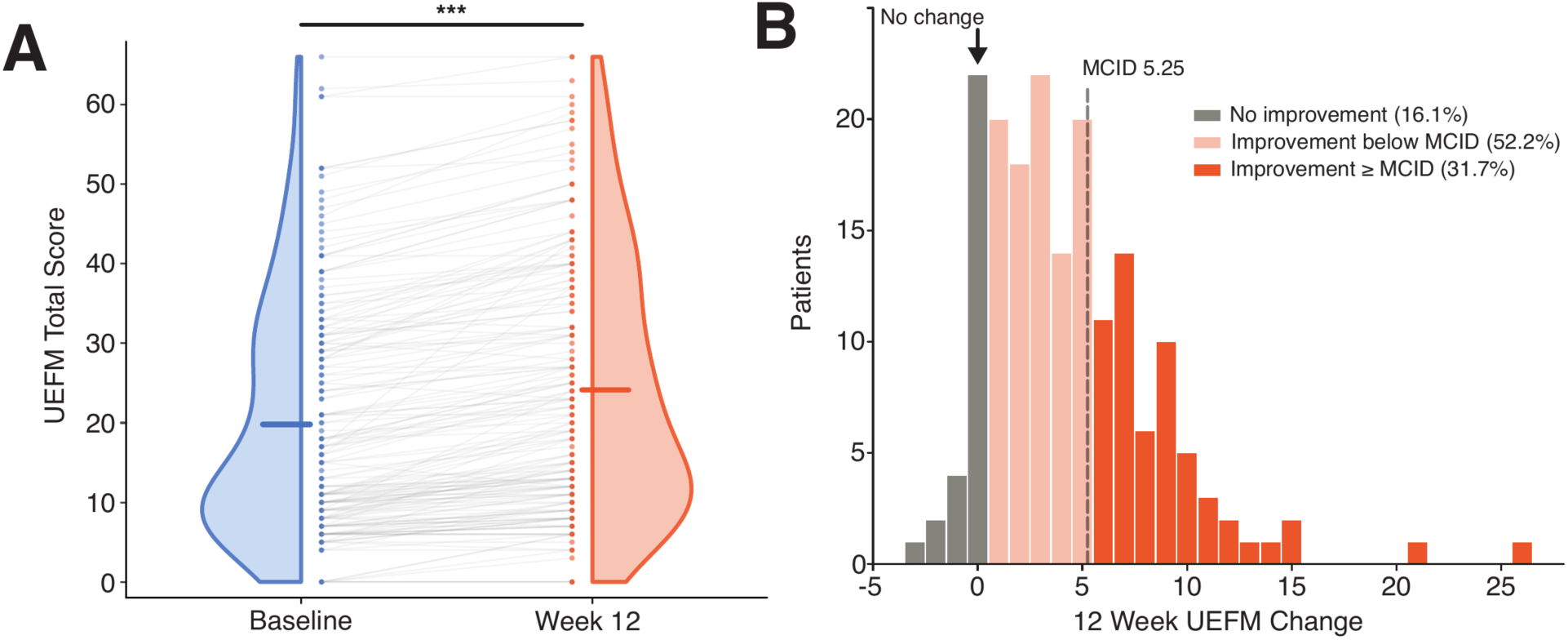
Changes in UEFM after 12 weeks of real-world BCI utilization (n = 180). (**A**) Violin plot showing paired changes in UEFM scores between baseline and week 12 (paired t-test, *** indicates *p* < 0.001). Horizontal bars show the mean of each distribution. (**B**) Histogram showing the distribution of observed UEFM changes at week 12 across patients.

Supplemental Table 4 compares the 180 patients with a week-12 assessment and the 174 patients without one, specifically, those with a 12-week assessment due but not recorded. Patients with and without a week-12 assessment showed no significant differences in baseline UEFM, age, time since stroke, or SDI.

### UEFM changes over one year

Observed UEFM changes significantly increased from baseline across all follow-up assessments, and improvements continued after week 12 (paired t-tests, all BH adjusted p < 0.01): +2.4 (SD 3.6; n = 269) at week 4, +3.5 (SD 3.8; n = 223) at week 8, +4.3 (SD 4.1; n=180) at week 12, +5.6 (SD 4.9; n = 149) at week 16, +6.0 (SD 5.2; n = 121) at week 20, +6.7 (SD 5.2; n = 92) at week 24, +6.5 (SD 5.3; n = 69) at week 28, +7.3 (SD 6.4; n = 52) at week 32, +7.2 (SD 6.7; n = 42) at week 36, +6.4 (SD 6.2; n = 19) at week 40, +9.3 (SD 10.3; n = 16) at week 44, +7.1 (SD 4.7; n = 10) at week 48, and +11.5 (SD 12.4; n = 15) at week 52. The full set of observed UEFM changes across all follow-up dates is shown in Supplemental Figure 2. Observed changes in UEFM for weeks 16 and 24 appear in Supplemental Figures 3 and 4, respectively. A comparison of baseline characteristics between the 369 patients with at least one follow-up assessment and the 83 patients with only a baseline assessment appears in Supplemental Table 4. Patients with vs. without follow-up assessments showed no significant differences in baseline UEFM, age, time since stroke, or SDI.

Due to data sparsity at later follow-up dates and missing assessments, we estimated mean UEFM change from baseline with an MMRM, which used all available assessments and within-patient correlations to estimate UEFM scores. The baseline coefficient was 1.047 (95% CI [1.022, 1.072]). Least-squares mean change from baseline was +4.2 (95% CI [3.8, 4.7]) at week 12, exceeded MCID at week 16 (+5.3; 95% CI [4.8, 5.9]), and demonstrated continued improvement up to 52 weeks (+8.1; 95% CI [6.8, 9.4]). The MMRM UEFM change trajectory is shown in Figure 3.

**Figure 3.**
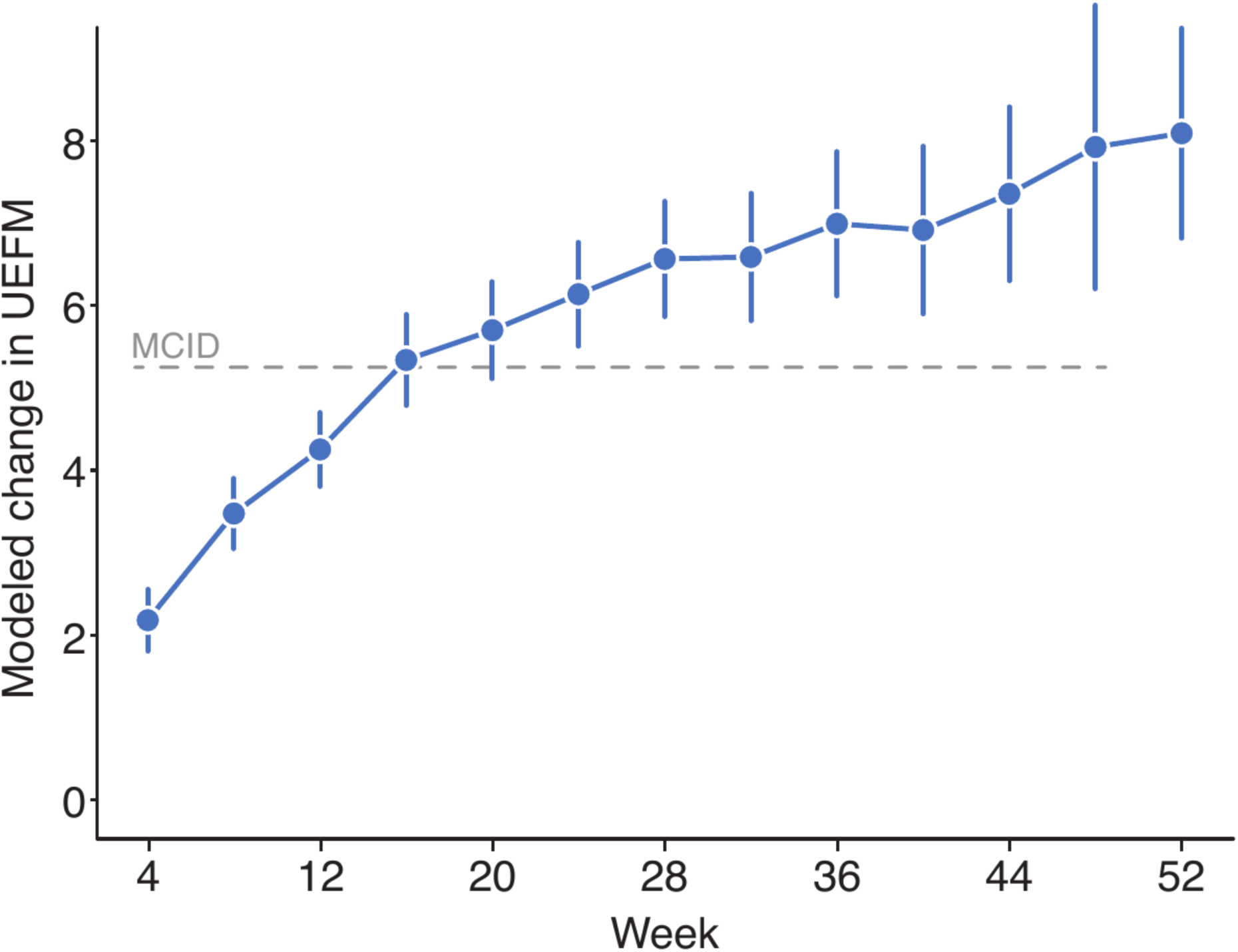
Change in UEFM over one year. Least-squares mean UEFM change from baseline, estimated with a mixed model for repeated measures (MMRM). Vertical bars represent 95% confidence intervals.

This pattern of continued improvement beyond week 12 persisted using the more conservative carry-forward multiple imputation approach. The least-squares mean change was +3.7 (95% CI [3.2, 4.3]) at week 12, exceeded MCID at week 28 (+5.3; 95% CI [4.3, 6.3]), and was +6.7 (95% CI [4.5, 8.9]) at week 52 (Supplemental Figure 5). Tipping point analysis determined that the estimated UEFM change from week 12 to week 52 remained above 0 up to a penalty (δ) of 2.5 (+1.5; 95% CI [0.1, 2.8]). The CI of the change from week 12 to week 52 included 0 at a δ of 3.0 points. A visualization of investigated δ is shown in Supplemental Figure 6. Across the observed means, the mixed model, and both sensitivity analyses, improvement continued beyond week 12.

In an exploratory analysis of 167 patients with baseline UEFM of 20-50 (167/452, 36.9%; mean baseline UEFM 32.0, SD 8.7), UEFM increased significantly from baseline at assessment weeks 12 (n = 67; 44.8% improved by at least MCID; mean UEFM change +5.3, SD 3.8, paired t-test p < 0.001), 16 (n = 52; 55.8% improved by at least MCID; mean UEFM change +6.6, SD 4.7, paired t-test p < 0.001), and 24 (n = 36; 75.0% improved by at least MCID; mean UEFM change +9.2, SD 4.8, paired t-test p < 0.001).

## Discussion

This study provides a real-world observational characterization of an at-home, noninvasive BCI therapy for upper extremity motor recovery after chronic stroke. Among patients with a paired assessment, mean UEFM improved by 4.3 points from baseline to week 12, with 31.7% of patients reaching MCID. Improvement did not plateau after 12 weeks. Both observed and model-estimated mean UEFM change exceeded MCID by week 16 and continued to increase. Together, these findings support the potential of at-home BCI therapy for upper-extremity motor rehabilitation after chronic stroke in real-world settings.

As an emerging technology, therapeutic BCI systems provide an alternative modality to conventional interventions. Users are required to dedicate cognitive effort and personal time to receive potential benefits from BCI use. As such, real-world evidence is valuable for assessing the potential for BCI adoption in routine care, especially in the context of debilitating pathologies affecting quality of life such as motor impairment following stroke. Currently, access to sustained rehabilitation remains a barrier for many survivors, particularly for those that depend on at-home solutions.^12,15^ While clinical trials are typically conducted at academic centers, with recruitment, oversight, and adherence incentives, the patients in this cohort self-administered BCI sessions at home under routine care. Understanding whether previously documented therapeutic outcomes from BCI therapies translate to real-world settings not only reinforces clinical findings but also provides quantitative evidence to support patient and provider expectations. The evidence provided in this study, while observational, suggests that real-world use of BCIs provides a promising and accessible option for chronic stroke survivors.

A defining feature of this study population is its motor impairment profile. Most patients (73.7%) had severe upper-extremity motor impairments at baseline. Greater baseline motor impairment is generally associated with limited recovery,^50,51^ and the magnitude of measured motor recovery varies with baseline severity.^37,52^ Despite the high proportion of severely impaired patients, a mean UEFM improvement exceeding MCID was observed by week 16, and this continued to increase out to one year. However, patients that reached MCID at weeks 12, 16, and 24 were significantly less impaired at baseline than patients that did not. It is possible that the mean week-12 improvement below MCID partly reflects the severity profile of this cohort, and that more severely impaired patients require a longer duration of BCI therapy to reach clinically meaningful improvement. Consistent with this possibility, patients with a baseline UEFM of 20-50, the inclusion range of previous clinical trials and inclusion recommendations,^44–47^ showed larger mean improvements than the full cohort at weeks 12, 16, and 24, with the mean improvement exceeding MCID by week 12. Future work evaluating how the rate, magnitude, and shape of improvement trajectories vary with baseline severity in a baseline UEFM-balanced cohort will be necessary to determine its association with at-home BCI-mediated motor recovery.

In addition to severe motor impairment, our cohort had a mean stroke chronicity (time from stroke onset to baseline assessment) of 5.4 years. Typically, in long-term stroke survivors, function and quality of life tend to decline rather than improve over longer intervals.^53–55^ Instead, our results suggest that meaningful clinical improvements may be possible in the chronic stage after stroke. Further, this chronicity profile and the observed improvements in UEFM resemble those reported in the BCI-REHAB randomized trial, reinforcing previous findings of meaningful improvements in the chronic stage after BCI intervention in a substantial fraction of subjects.^25^

Compared to previously reported UEFM outcomes in prospective and randomized clinical trials, the mean week-12 improvement in our cohort was smaller.^24,25^ At later timepoints, however, our observed and model-estimated improvements from baseline were comparable to the range reported by those trials, with the observed mean improvement reaching +5.6 by week 16 and +6.7 by week 24. It is possible that comparable gains appearing later are consistent with slower accrual of therapeutic benefits under the variable conditions of routine care. For example, greater scheduled therapy time has been associated with larger motor gains after stroke.^9^ The BCI-REHAB trial instructed patients to complete five one-hour sessions per week.

In contrast, our cohort used the BCI for an average of 3.6 sessions per week, and we did not analyze session duration or other utilization metrics. It is possible that degree of the UEFM improvement observed in this study is associated with extent of BCI utilization. Future work characterizing and examining the relationship between BCI utilization and UEFM improvements will be necessary to establish how much use is required for clinically meaningful improvement and to inform dosing recommendations in routine care. Further, based on the characterized improvements from this study, future study designs, or recommendations for at-home BCI-mediated upper-extremity motor recovery should consider evaluating primary outcome dates beyond 12-weeks, as the present evidence suggests that improvement may continue past previously evaluated outcome dates.

Taken together with previous real-world evidence,^29^ these findings support at-home BCI therapy as a treatment where upper-extremity improvements can emerge during routine care, not only in controlled trials. To our knowledge, this is the first characterization of therapeutic outcomes at this scale in prescribed users of an FDA-cleared BCI, analyzed under a pre-specified plan, and followed from the start of therapy to one year. Understanding how novel BCI therapies perform as a rehabilitation option in real-world settings provides patients and providers with evidence to set realistic expectations and judge the therapy for routine care. Notably, our results provide an encouraging signal for the future of BCI therapies for stroke. Within our cohort, we found that clinically meaningful improvements in motor function can occur years after stroke, even in patients whose recovery is often assumed to have plateaued. Although this study is observational, it establishes an initial real-world evidence base for larger, future prospective controlled trials that can clarify the efficacy and target population for BCI adoption into routine stroke care.

### Limitations

This study has several limitations. It was observational and single-arm, without a randomized comparator. Therefore, the within-cohort change and the outcome characterizations are contextual; extent of rehabilitation therapy outside of study procedures was not measured and might have impacted the observed change over time, though such therapy is typically absent in patients with chronic stroke.^56^ The UEFM was administered by unblinded occupational therapists remotely by video conference. While video administration of the Fugl-Meyer shows adequate agreement with in-person administration,^57,58^ it does not remove the expectation effects that may accompany unblinded assessment.^59^ BCI therapy was self-administered at home, and the analysis could not document or correct for environmental factors that may have influenced therapy delivery, such as reliable internet access,^60^ correct headset placement and upkeep, and adherence to the intended BCI task. Further, this study cohort consists of patients engaged with routine care, which includes routine follow-up assessments. As reflected in our cohort profile in Figure 1B, many real-world BCI users do not receive UEFM assessments, and therefore, our findings describe only patients who receive clinical follow-up services. Further, only patients with outcome assessments contributed to the analyses. If unassessed patients improved less, the estimates overstate improvement. Finally, this analysis did not model predictors of outcome, and future work should examine how recovery varies with age, sex, socioeconomic status, and stroke pathology.

## Acknowledgments

Anthropic’s Claude and Claude Code were used for writing and editing assistance; their use is described in the Methods (AI use).

