## Supplemental Materials for "Real-World Improvement in Motor Status Among Regular Users of a Home-Based Brain-Computer Interface After Stroke"

### Supplemental Material

A Retrospective Real-World Assessment of Motor Status After Stroke Using a Home-Based Brain-Computer Interface

#### Contents

Supplemental Methods: Statistical Analysis Plan, Version 1.0 (signature page omitted) p. 2

Supplemental Figures 1–6 p. 15

Supplemental Tables 1–4 p. 21

STROBE Statement: checklist for cohort studies p. 25

Page numbers refer to this PDF. Checklist page and line numbers refer to the line-numbered manuscript.

Docusign Envelope ID: 601441CA-296E-8E2E-81A6-286A9EAF73CF



**Study title.** A Real-World Assessment of Post-Stroke Motor Function Improvements Using IpsiHand®

**Approved by.** The study team (primary analyst, study statistician, and others involved in the data processes), signing below.

> **Pre-specification.** This plan fixes the analysis methods before the final analysis dataset is locked, so that the reported characterization rests on pre-specified methods. Any deviation will be recorded in the Version History.

CONFIDENTIAL / PROPRIETARY: This document contains sensitive information. Unauthorized copying, distribution, or disclosure is strictly prohibited.

Docusign Envelope ID: 601441CA-296E-8E2E-81A6-286A9EAF73CF

**Table of Contents**

Signatures ... 2
Version History ... 4
Abbreviations ... 5
1 Introduction ... 6
2 Roles and responsibilities ... 6
3 Study overview ... 7
3.1 Endpoint and estimands ... 7
3.2 Study description ... 7
4 Analysis population and data ... 8
4.1 Full Analysis Set (FAS) population: ... 8
4.2 Per Protocol Population 1- Device Use Compliance (PPP-1_DUC) ... 8
4.2.1 Inclusion and exclusion criteria ... 8
4.2.2 Analysis dataset and data management ... 9
4.2.3 Outcome, timepoints, and definitions ... 9
5 Analysis ... 10
5.1 General Considerations ... 10
5.2 Subpopulation-Cohort characteristics ... 10
5.3 Endpoint ... 11
5.3.1 Defining continued improvement ... 11
5.3.2 Mixed model for repeated measures (MMRM) ... 11
5.4 Treatment of missing values ... 12
5.5 Sensitivity analyses ... 12
6 Analysis software and reproducibility ... 13
7 Validation of analyses ... 13
8 Reporting and STROBE adherence ... 13
9 Changes to the analysis plan ... 13
10 References ... 13

CONFIDENTIAL / PROPRIETARY: This document contains sensitive information. Unauthorized copying, distribution, or disclosure is strictly prohibited.

Docusign Envelope ID: 601441CA-296E-8E2E-81A6-286A9EAF73CF

**Version History**

| Version Number | Effective Date | Reason for Change |
|---|---|---|
| 0.1 | July 28, 2026 | First Draft |
| 1.0 | August 17th, 2026 | Final Draft |

CONFIDENTIAL / PROPRIETARY: This document contains sensitive information. Unauthorized copying, distribution, or disclosure is strictly prohibited.

**Abbreviations**

| Term | Definition |
|---|---|
| BCI | Brain–computer interface |
| BIC | Bayesian Information Criterion |
| CI | Confidence interval |
| ICH E9(R1) | International Council for Harmonisation E9(R1) addendum on estimands and sensitivity analysis |
| LMCF | Last mean carried forward |
| LOCF | Last observation carried forward |
| LS mean | Least-squares mean |
| MAR | Missing at random |
| MCAR | Missing completely at random |
| MCID | Minimal clinically important difference |
| MI | Multiple imputation |
| ML / REML | Maximum likelihood / restricted maximum likelihood |
| MMRM | Mixed model for repeated measures |
| MNAR | Missing not at random |
| RWE | Real-world evidence |
| STROBE | Strengthening the Reporting of Observational Studies in Epidemiology |
| UEFM | Upper Extremity Fugl-Meyer assessment (Unit Measure 0–66) |

CONFIDENTIAL / PROPRIETARY: This document contains sensitive information. Unauthorized copying, distribution, or disclosure is strictly prohibited.

Docusign Envelope ID: 601441CA-296E-8E2E-81A6-286A9EAF73CF

#### 1 Introduction

This Statistical Analysis Plan (SAP) specifies the analysis of upper-extremity motor recovery in a real-world cohort of stroke survivors who used the IpsiHand® system, a fully remote home-based brain–computer-interface (BCI) therapy authorized for chronic stroke-induced upper-limb impairment. The primary outcome is the Upper Extremity Fugl-Meyer assessment (UEFM), measured over the course of routine device use.

This study presents a retrospective observational characterization of the changes in UEFM within a cohort of real-world users. Each patient in the cohort utilized IpsiHand® as part of the Kandu Medical Services clinical program, and there is no randomized comparator. The analysis therefore characterizes how UEFM changes within this population over time.

This document defines the populations and the study procedures, the outcome and timepoints, the analysis of the endpoint, the handling of missing data, the sample size and precision, the analysis software, the validation of results, and the reporting standard. It follows the reporting expectations of the STROBE statement for observational cohort studies (Section 8).

#### 2 Roles and responsibilities

| Name | Role | Responsibility |
|---|---|---|
| Seth Wilk, PhD | Principle investigator | Lead and supervise the realization of the study, review data and results |
| Gregory E. Wilding, PhD | Independent validation analyst | Advise development of statistical analysis plan and performance of validation analysis |
| Steven C. Cramer, MD | Primary author | Lead the practical and scientific contributions to the study |
| Phillip Demarest, PhD | Primary analyst | Lead the creation of the statistical analysis plan, review and maintain data, perform monthly analysis of the full analysis, perform final study analysis |

The primary analyses will be implemented by the primary analyst and verified by the independent validation analyst before results are considered final.

CONFIDENTIAL / PROPRIETARY: This document contains sensitive information. Unauthorized copying, distribution, or disclosure is strictly prohibited.

Docusign Envelope ID: 601441CA-296E-8E2E-81A6-286A9EAF73CF

#### 3 Study overview

##### 3.1 Endpoint and estimands

The analysis addresses two aims, both describing change within the treated cohort over time:

**Enpoint.** Characterize the changes in UEFM from baseline to subsequent scores in IpsiHand® users.

The endpoint aims to determine whether changes in UEFM are detected within a real-world cohort and characterize such changes.

Following the International Council on Harmonisation of Technical Requirements for Pharmaceuticals for Human Use (2019) framing, the estimand for both aims is a within-cohort quantity:

- **Population:** real-world IpsiHand® users meeting the criteria of Section 4.1.
- **Treatment condition:** The IpsiHand® System is an FDA-cleared, non-invasive therapy for chronic stroke-induced upper limb motor impairment that uses a brain-computer interface (BCI) to translate intent-to-move signals into hand movement." Patients were "prescribed IpsiHand® and completed regular UEFM assessments as part of routine care.
- **Endpoint:** Patient UEFM total score, assessed at baseline and at clinical care engagement post introduction of device use
- **Population-level summary:** the within-cohort mean change from baseline to any subsequent follow up timepoint, estimated as the model-based contrast between the one-year and 12-week outcome measurements.
- **Intercurrent events and strategies:** There are no pre-defined intercurrent events. Missing values will be assumed missing at random and handled via direct likelihood as the primary assumption, with sensitivity analyses conducted.

These estimands are interpreted as descriptions of the real-world cohort's trajectory over time.

##### 3.2 Study description

The cohort is drawn from the Kandu clinical program for IpsiHand ®. As there is no separate study protocol, the relevant procedures are described here.

- **Identification and enrollment.** Patients were prescribed IpsiHand® in accordance with its FDA-cleared indication and used it as part of routine care. The population comprises the users who consented to share their demographic information and outcomes and who meet the criteria of Section 4.1.
- **Assessment schedule.** UEFM is assessed on a nominal visit schedule over the course of device use every 4 weeks. Not every patient is assessed at every scheduled visit and observed assessment dates scatter around the nominal week. Due to variation in patient schedules, not all assessments were completed on the appropriate corresponding nominal date. Variation in

CONFIDENTIAL / PROPRIETARY: This document contains sensitive information. Unauthorized copying, distribution, or disclosure is strictly prohibited.

observed assessment dates will be quantified and reported. Assessment was conducted via video conference by unblinded occupational therapists trained to perform UEFM assessments.

- **Baseline.** Baseline is the total UEFM score measured prior to initial prescription and utilization of the IpsiHand® device.
- **Device provision.** IpsiHand® is a home-based device provided on prescription in accordance with its FDA cleared indication.
- **Assessment context.** Assessments are collected as part of routine care, with no randomization or masking; UEFM assessors are aware of the therapeutic context. Any resulting assessment bias will be noted as a limitation (Section 8).

#### 4 Analysis population and data

Full analysis and summary statistics will be used to characterize the full analysis set (FAS), according to the details outlined in the study protocol. The FAS includes all persons prescribed IpsiHand® from 01/01/2023 until 08/21/2026 with document of system receipt.

##### 4.1 Full Analysis Set (FAS) population:

Inclusion Criteria:

(1) Age 18 years and older
(2) Prescribed IpsiHand® by a provider

##### 4.2 Per Protocol Population 1- Device Use Compliance (PPP-1_DUC)

###### 4.2.1 Inclusion and exclusion criteria

Inclusion and exclusion criteria are defined by a combination of program eligibility and analysis considerations. Eligibility follows the IpsiHand® FDA-authorized indication (FDA De Novo DEN200046; 21 CFR 890.5420). These considerations include age, stroke chronicity, and stroke clinical manifestation. Analysis considerations include criteria that best enable characterization of primary outcomes in a real-world user cohort of the IpsiHand® device. These considerations include usage of the device, availability of outcome measures, and exclusion of major clinical events unrelated to device utilization.

Inclusion:

(1) Age 18 years and older Prescribed IpsiHand® by a provider in accordance with the current FDA cleared indication
(2) Stroke sustained at least six months before initial IpsiHand® use
(3) Presence of hemiparesis or hemiplegia as measured by cognitive function, language, motor function or visual attention assessments

CONFIDENTIAL / PROPRIETARY: This document contains sensitive information. Unauthorized copying, distribution, or disclosure is strictly prohibited.

Docusign Envelope ID: 601441CA-296E-8E2E-81A6-286A9EAF73CF

Exclusion:

(1) Absence of a baseline UEFM measurement
(2) Failure to use the device for an average of at least one session per week
(3) Any major detrimental event after starting IpsiHand® usage (a significant medical event, injury, or change in neurological or functional status that may compromise the patient's safety or ability to continue IpsiHand® therapy, for example, recurrent stroke, hospitalization, fall with injury, fracture, or surgery)

**4.2.2 Analysis dataset and data management**

The analysis will operate on a single de-identified dataset assembled from the routine-care records of the Kandu Medical Systems, with one patient per stable de-identified account identifier. As such, the dataset exists in a live refreshable export. The dataset used in each analysis will be frozen for final analysis on the date indicated in a report. Data will be formatted and exported in tabular format. Only the variables outlined within this SAP will be exported as part of the dataset used in the study.

**4.2.3 Outcome, timepoints, and definitions**

**Primary outcomes.** UEFM total score (range 0–66) treated as a continuous bounded outcome for paired comparisons, and change in UEFM relative to baseline as the endpoint in our statistical models.

**MCID.** The minimal clinically important difference is defined as 5.25 points, within the 4.25–7.25 anchor-based range reported by Page et al. (2012, PMID 22282773). Baseline UEFM will be grouped into severity bands (severe 0–28, moderate 29–42, mild 43–66) for description.

**Baseline.** Baseline is the UEFM total score recorded on or before the date of initial IpsiHand® use, within 30 days prior to that date.

**Age.** Age is defined at baseline (years elapsed from date of birth to the baseline date).

**Timepoints and windowing.** UEFM will be analyzed on nominal scheduled weeks and binned within every 4 weeks +/- 2 weeks (baseline, 4, 8, 12, 16, 20, 24, 28, 32, 36, 40, 44, 48 and 52) with baseline as the origin and the nominal week as the time axis. Variance within each nominal week will be reported as a range.

**Unit of analysis.** The patient is the unit of analysis. Each patient contributes repeated UEFM measurements across timepoints. The paired analyses (Section 5.2) will operate on within-patient change on complete pairs; the longitudinal models (Section 5.3) will use each patient's available repeated measurements.

CONFIDENTIAL / PROPRIETARY: This document contains sensitive information. Unauthorized copying, distribution, or disclosure is strictly prohibited.

Docusign Envelope ID: 601441CA-296E-8E2E-81A6-286A9EAF73CF

**5 Analysis**

Analysis of the FAS will be conducted monthly as part of the study protocol to ensure full summarization of the study population. Characteristics including population size, statistical summaries of demographic information, and statistical summaries of outcome measures will be reported according to the inclusion criteria of the FAS. Continuous characteristics will be summarized by mean and standard deviation. Categorical characteristics by count and percentage. Characterization of the FAS will include summarization of: age, sex, UEFM handedness, socioeconomic status, affected limb, and time since onset of stroke. Monthly analysis will be conducted by the primary analyst.

**5.1 General Considerations**

The pre-specified two-sided significance level for all hypothesis testing is α = 0.05. P-values are two-sided and will be reported to three significant figures or as "<0.001" when smaller

Multiplicity: Comparative analysis subject to multiple corrections will be accounted for using a Benjamini-Hochberg procedure to control for false-discovery rate.

Descriptive summaries will report the sample size relevant to each calculation and the number of missing observations. Continuous summaries will carry one more decimal place than the source data; percentages will be reported to one decimal place.
Change in UEFM will be calculated as:

For patient *i* at nominal week *t*, the change from baseline in UEFM total score is

$$\Delta_{\{it\}} = Y_{\{it\}} - Y_{\{i0\}}$$

where $Y_{\{it\}}$ is the UEFM total score at nominal week *t* and $Y_{\{i0\}}$is that patient's baseline score.

Reporting will follow the STROBE cohort checklist (Section 8).

**5.2 Subpopulation-Cohort characteristics**

The subpopulation will be summarized in a characteristics table, overall and by baseline severity band: age, sex, baseline UEFM, enrollment status (whether the patient is still actively using the IpsiHand® system or has completed therapy at the time of data freezing), time since onset of stroke, affected upper extremity, handedness, and socioeconomic status. Continuous characteristics will be summarized by mean, standard deviation, median, interquartile range, and range with the observed n; categorical characteristics by count and percentage. Subpopulation characteristics will be compared descriptively across severity bands; no formal subgroup hypothesis test is planned.

CONFIDENTIAL / PROPRIETARY: This document contains sensitive information. Unauthorized copying, distribution, or disclosure is strictly prohibited.

##### 5.3 Endpoint

The endpoint will estimate the within-patient change in UEFM from baseline to any follow up timepoint. This descriptive analysis is centered on the mean paired change:

- **Paired t-test** of the within-patient change, reporting the mean change with its two-sided 95% confidence interval.
- **Bootstrap 95% confidence interval** for the mean change (patient-level resampling), reported along the t-interval.
- **Distribution summary:** the mean and standard deviation, the median and interquartile range, and a standardized paired effect size (Cohen's d).
- **Responder summaries:** the percentage improving (change > 0) and the percentage reaching the MCID.

###### 5.3.1 Defining continued improvement

The characterization will distinguish three trends following 12 weeks of therapy, read from the later-phase parameters of the model below:

- **Continued improvement:** mean UEFM continues to increase after week 12 and the one-year (mean exceeds the 12-week mean).
- **Maintenance:** Mean UEFM after week 12 is approximately equal to the 12-week mean.
- **Regression:** Mean UEFM declines after week 12 (the one-year mean falls below the 12-week mean).

This is a population-average characterization over the whole cohort; it describes the cohort's mean trajectory and does not condition on whether individual patients improved before week 12.

###### 5.3.2 Mixed model for repeated measures (MMRM)

In order to characterizes changes in UEFM after 12 weeks, we will use a mixed model for repeated measures (MMRM), with time entered as an unordered categorical factor. Each scheduled visit receives its own independently estimated mean change, so the model imposes no functional shape on the trajectory.

For patient *i* at nominal visit *t*, UEFM total score is modeled as

$$Y_{it} = \mu_t + \beta Y_{i0} + e_{it}$$

where $Y_{it}$ is patient I's baseline UEFM and the within patient residual vector is

$$\mathrm{e_i} = \left(\mathrm{e_{i1}}, \ldots, \mathrm{e_{iJ_i}}\right)^{\mathrm{T}} \sim \mathrm{N}(0, \Sigma_\mathrm{i})$$

Here, $\mu_t$ is the least-squares mean change from baseline at nominal visit *t*, entered as a categorical factor so that each visit's mean is free of any assumed shape. $J_i$ is the number of visits

CONFIDENTIAL / PROPRIETARY: This document contains sensitive information. Unauthorized copying, distribution, or disclosure is strictly prohibited.

Docusign Envelope ID: 601441CA-296E-8E2E-81A6-286A9EAF73CF

patient *i* contributed, and $\Sigma_i$ is a matrix of patient *i*'s observed visits, of a within-patient covariance $\Sigma$.

The within-patient covariance is unstructured across the nine visits as the primary specification. If the unstructured model does not converge or returns a non-positive definite estimate, we will implement a covariance replacement (for example, a heterogeneous Toeplitz; O'Kelly & Ratitch, 2014). Confidence intervals for the least-squares means and their contrasts will use the model-based repeated-measures covariance with a Kenward-Roger degrees-of-freedom adjustment (Kenward & Roger, 1997).

**5.4 Treatment of missing values**
Follow-up length varies across the cohort, so not every patient has a score at every timepoint. The primary analysis will assume data are missing at random and will handle missing values by direct likelihood (O'Kelly & Ratitch, 2014).

**5.5 Sensitivity analyses**
For sensitivity analysis, we will perform two approaches: a tipping point analysis to assess departures from the MAR assumption, and a multiple-imputation analysis under an alternative assumption for the unobserved values.

For tipping-point analysis, a fixed penalty $\delta$ will be subtracted from each imputed post-12-week value (shifting values against the direction of thee estimated improvement). $\Delta$ will be increased until the model conclusion no longer holds. This analysis will report the smalles $\delta$ (the departure from MAR) that overturns the conclusion (Permutt, 2016).

Multiple imputation will be conducted by imputing each missing value from the patient's last observed value with an added stochastic draw, across M imputed datasets combined by Rubin's rules (Carpenter, Roger & Kenward, 2013). We will use 50 imputed datasets, where each completed dataset will be analyzed using the MMRM model, and estimates will be combined by Rubin's rules. The multiple-imputation estimate of the change after week 12, and of the 12-week change will be reported alongside the primary analysis's estimate. This method will support assessment of the continued-improvement conclusion under a more conservative assumption than MAR.

Baseline UEFM of the patients observed in exploratory timepoints, such as Week 16 and post-week 16 may be characterized in similar analyses.

The baseline UEFM of the patients observed at one year will be compared with that of the rest of the cohort (Mann-Whitney U test) to characterize whether the one-year subset is representative; the direction of any difference will be used to read the observed one-year change conservatively or with discount, as appropriate.

CONFIDENTIAL / PROPRIETARY: This document contains sensitive information. Unauthorized copying, distribution, or disclosure is strictly prohibited.

**6 Analysis software and reproducibility**

Primary analyses will be implemented in Python (version 3.9 or higher): statsmodels for the mixed models, scipy.stats for the nonparametric tests, and numpy / pandas for data handling. The categorical-time MMRM (Model B) may additionally be fit in R (for example the mmrm or nlme package) where its covariance handling is preferable. The statistical validation will conducted in SAS. Random seeds (for example in a bootstrap or a multiple-imputation draw) will be fixed and documented for reproducibility. Analysis code will be made available upon request.

**7 Validation of analyses**

Independent statistical validation

As an additional layer of methodological rigor, the de-identified per-protocol analysis dataset and the analysis pipeline will be forwarded to the independent validation analyst (Gregory Wilding), who will independently replicate and confirm the primary results. Any discrepancy between the primary and the independent analyses will be investigated and reconciled before the results are considered final, and the validation will be documented. Analysis of the FAS will not be conducted by the independent validation analyst.

**8 Reporting and STROBE adherence**

The manuscript will be reported in accordance with the STROBE statement for observational cohort studies (von Elm et al., 2007; PMID 18064739). A completed STROBE checklist will accompany the submission, mapping each item to its location in the manuscript. Study limitations, including the single-arm design without a randomized comparator and potential bias from unblinded assessment, will be reported under STROBE item 19.

**9 Changes to the analysis plan**

Changes made after the plan is finalized will be recorded in dated amendments, each stating the change and its rationale. A data error discovered after the analysis dataset is frozen will be documented in an errata record, and any correction applied in the analysis code will be approved and recorded before results are regenerated.


CONFIDENTIAL / PROPRIETARY: This document contains sensitive information. Unauthorized copying, distribution, or disclosure is strictly prohibited.

*NeuroEngineering and Rehabilitation*, 23(1), 68. (PMID 41566501.) — closest prior real-world IpsiHand® analysis (retrospective, n=56, benefit beyond 12 weeks).

- Rustamov, N., Souders, L., Sheehan, L., Carter, A., & Leuthardt, E.C. (2025). IpsiHand® brain-computer interface therapy induces broad upper extremity motor rehabilitation in chronic stroke. *Neurorehabilitation and Neural Repair*, 39(1), 74–86. (PMID 39345118.) — prospective IpsiHand® study (n=26, 12 weeks) at the established endpoint.
- International Council for Harmonisation. (2019). *E9(R1): Statistical principles for clinical trials — addendum: estimands and sensitivity analysis in clinical trials.*
- Kenward, M.G., & Roger, J.H. (1997). Small sample inference for fixed effects from restricted maximum likelihood. Biometrics, 53(3), 983–997. (PMID 9333350.) — Kenward-Roger degrees-of-freedom adjustment.
- O'Kelly, M., & Ratitch, B. (2014). Clinical trials with missing data: a guide for practitioners. Wiley. — direct-likelihood analysis under MAR and MMRM covariance structures.
- Carpenter, J.R., Roger, J.H., & Kenward, M.G. (2013). Analysis of longitudinal trials with protocol deviation: a framework for relevant, accessible assumptions, and inference via multiple imputation. *Journal of Biopharmaceutical Statistics*, 23(6), 1352–1371. (PMID 24138436.) — reference-based multiple imputation (LMCF family).
- Permutt, T. (2016). Sensitivity analysis for missing data in regulatory submissions. *Statistics in Medicine*, 35(17), 2876–2879. (PMID 26567763.) — tipping-point / delta-adjustment.
- Schwarz, G. (1978). Estimating the dimension of a model. *Annals of Statistics*, 6(2), 461–464. — BIC.
- von Elm, E., Altman, D.G., Egger, M., Pocock, S.J., Gøtzsche, P.C., & Vandenbroucke, J.P. (2007). The Strengthening the Reporting of Observational Studies in Epidemiology (STROBE) statement. *Lancet*, 370(9596), 1453–1457. (PMID 18064739.)

CONFIDENTIAL / PROPRIETARY: This document contains sensitive information. Unauthorized copying, distribution, or disclosure is strictly prohibited.

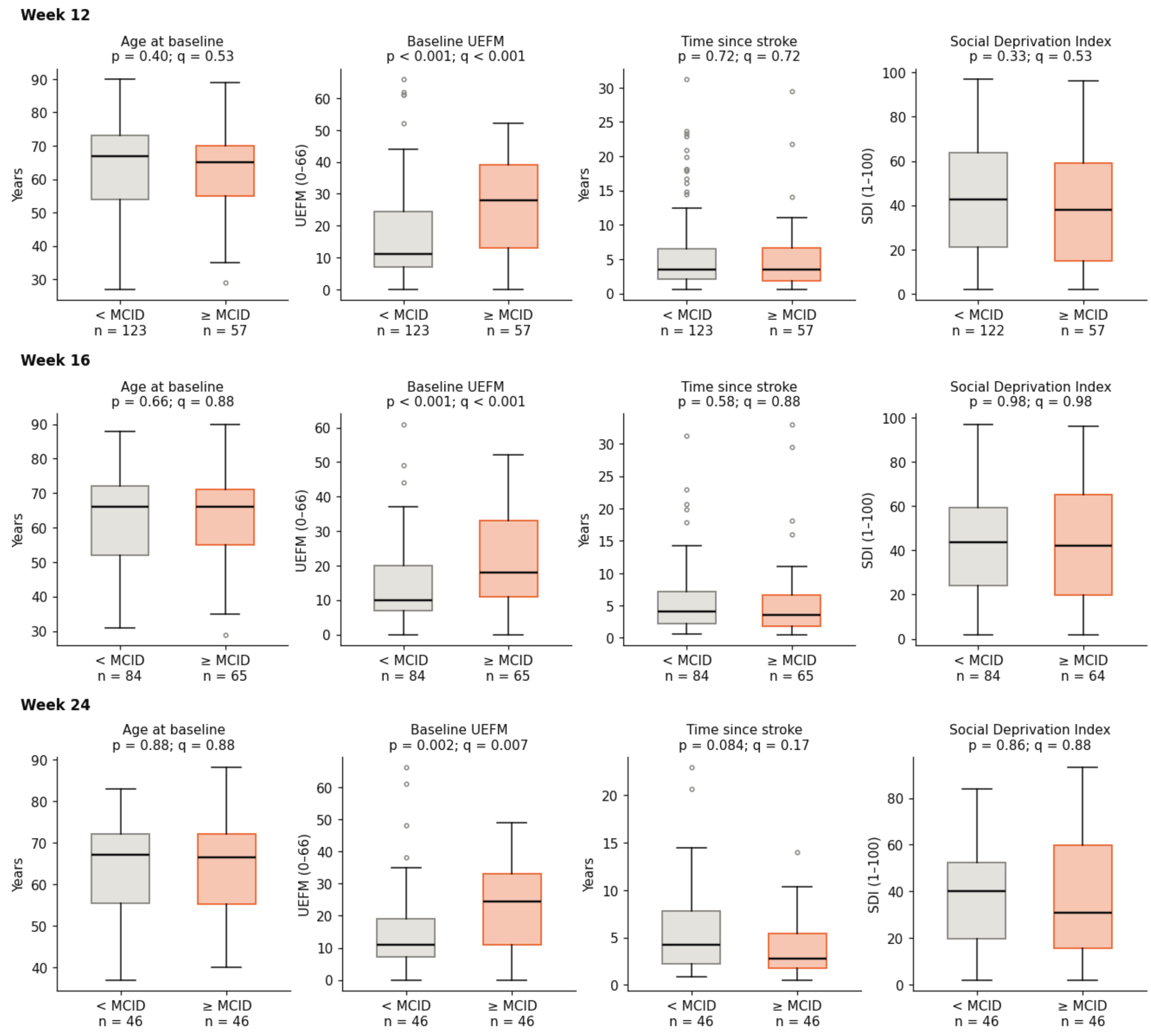


***Supplemental Figure 1.*** **Baseline characteristics of patients that reach MCID on weeks 12, 16, and 24.** Box plots comparing baseline characteristics (age, baseline UEFM, time since stroke, social deprivation index) of patients that reached MCID with those that did not at weeks 12, 16, and 24 (Wilcoxon rank-sum test, BH procedure adjusted p-values).

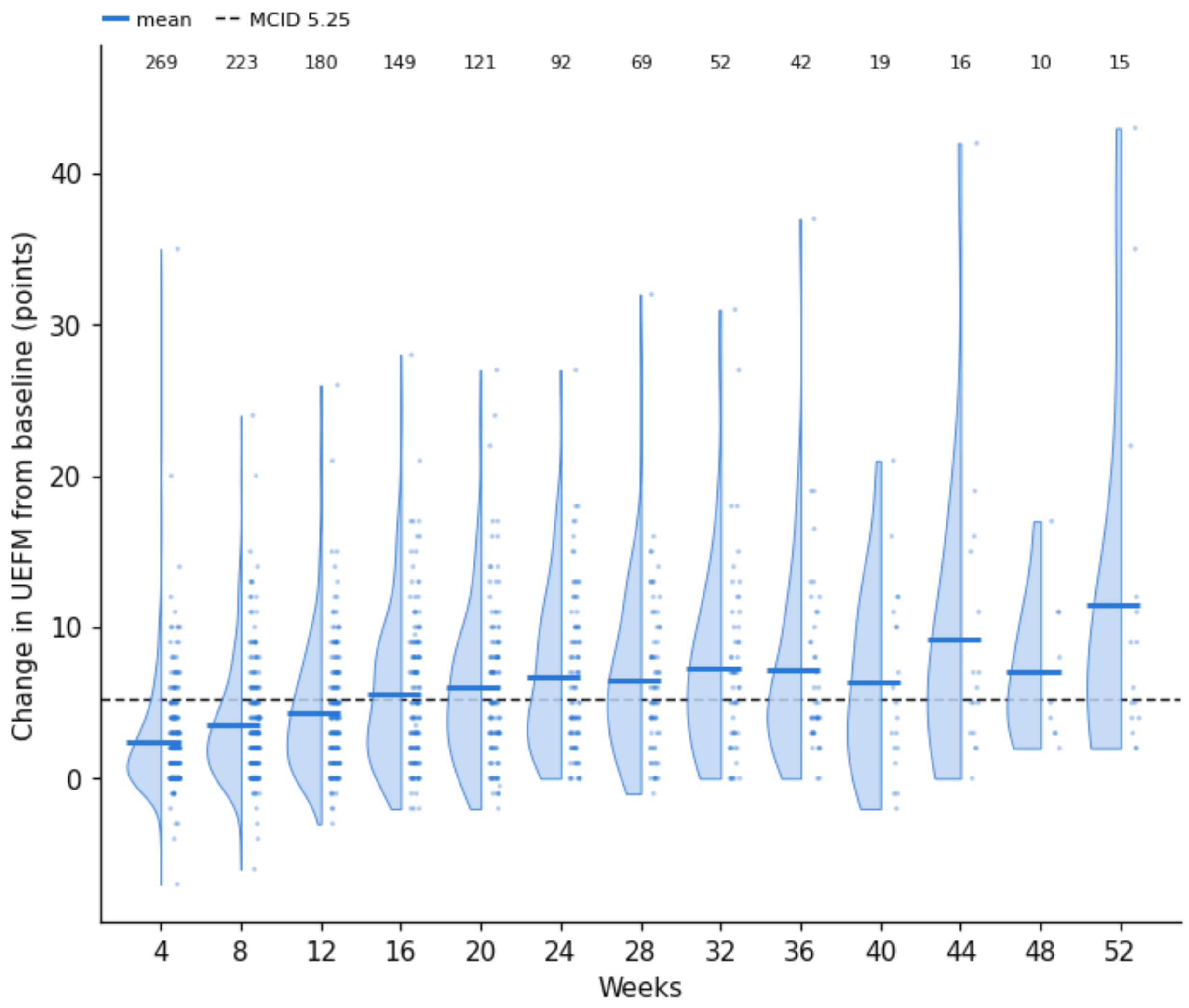


***Supplemental Figure 2.*** **Observed changes in UEFM.** Swarm plot and kernel densities showing changes in UEFM across all patients and follow up assessments. Horizontal bars show mean UEFM change. Numbers indicate observations at each follow-up.

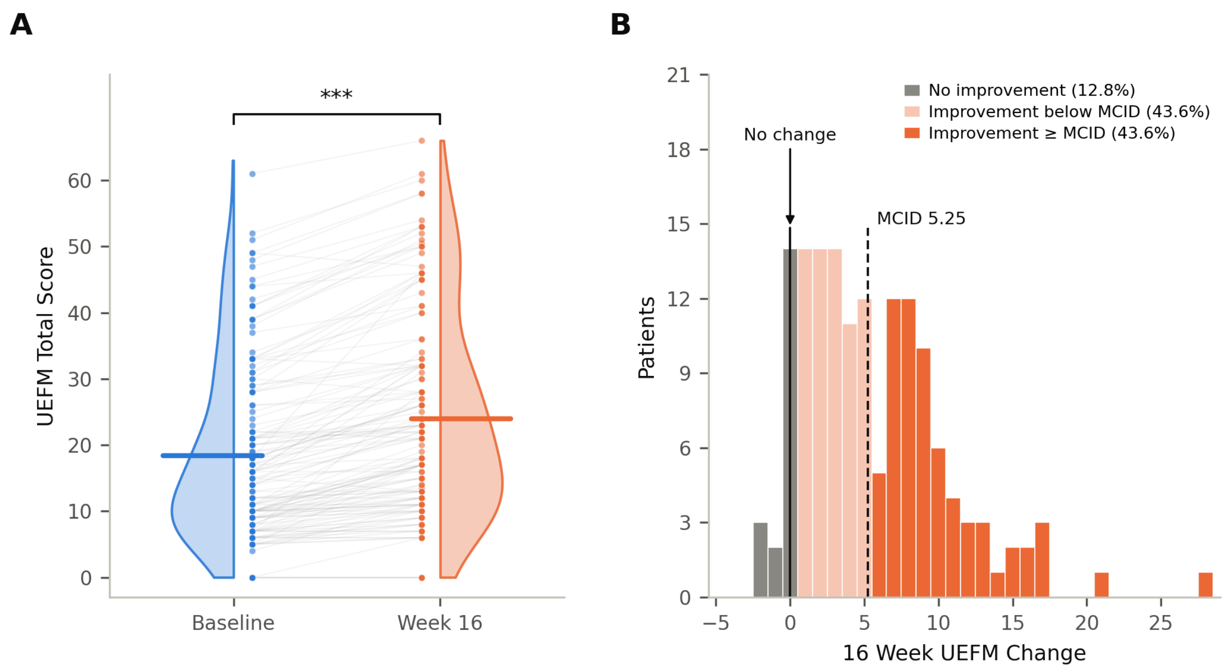


***Supplemental Figure 3.* Changes in UEFM after 16 weeks of real-world BCI utilization (n = 149).** (**A**) Violin plot showing paired changes in UEFM scores between baseline and week 16 (paired t-test, *** indicates $p < 0.001$). Horizontal bars show the mean of each distribution. (**B**) Histogram showing the distribution of observed UEFM changes at week 16 across patients.

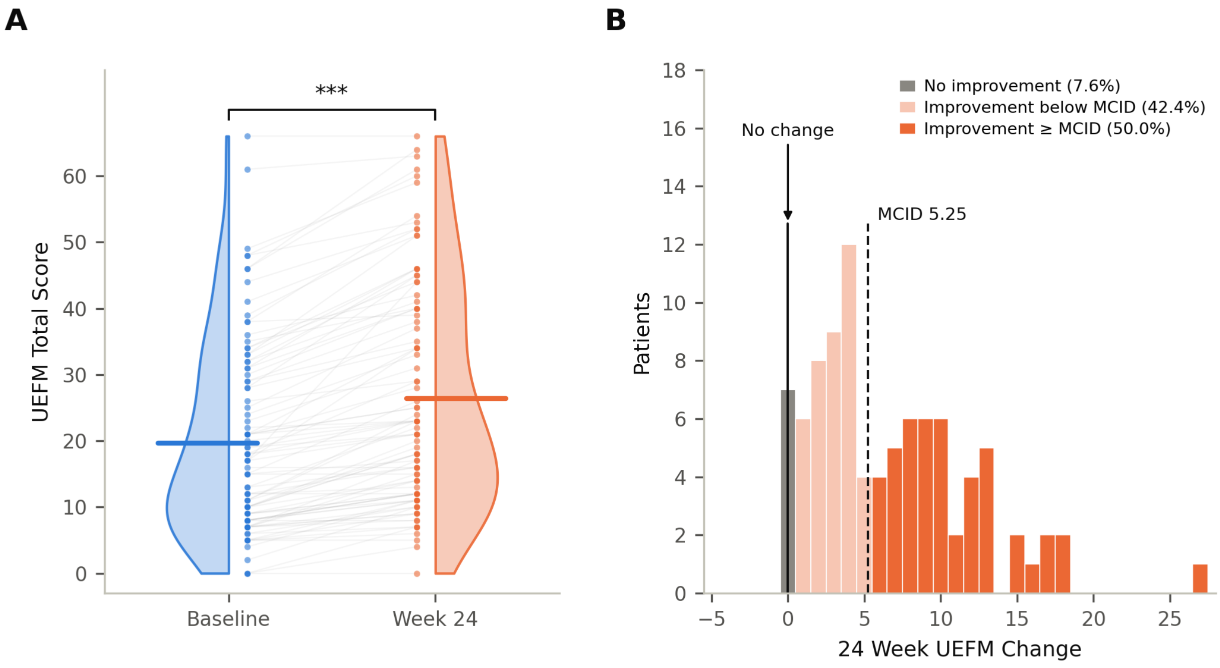


***Supplemental Figure 4.* Changes in UEFM after 24 weeks of real-world BCI utilization (n = 92).** (**A**) Violin plot showing paired changes in UEFM scores between baseline and week 24 (paired t-test, *** indicates $p < 0.001$). Horizontal bars show the mean of each distribution. (**B**) Histogram showing the distribution of observed UEFM changes at week 24 across patients.

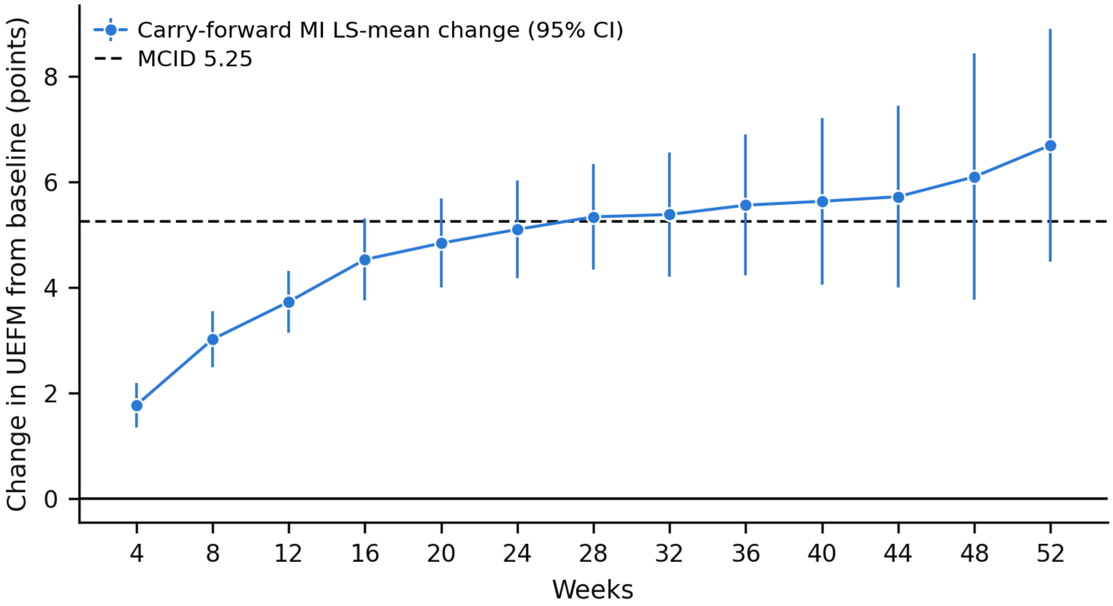


***Supplemental Figure 5.*** **Carry-forward multiple imputation LS mean change.** UEFM change trajectory under imputation using carry-forward multiple imputation.

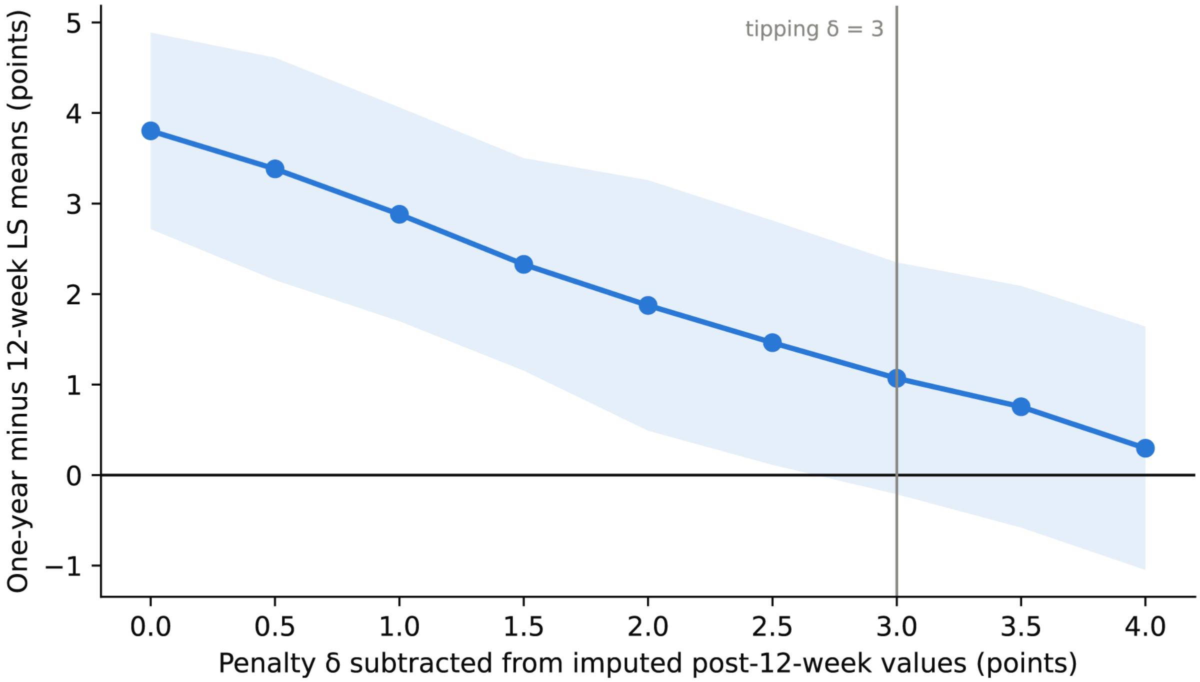


***Supplemental Figure 6.*** **Tipping point analysis.** Effect of penalty term δ on the estimated UEFM change from week 12 to week 52. Tipping point is defined as the minimum δ where 0 UEFM change falls within the 95% confidence interval.

***Supplemental Table 1.*** Follow-up UEFM availability.

| **Follow-up Week** | **# Observed** | **# Due but absent** | **# Not yet due** | **% Observed** |
|---|---|---|---|---|
| **4** | 269 | 100 | 0 | 72.9 |
| **8** | 223 | 130 | 16 | 63.2 |
| **12** | 180 | 160 | 29 | 52.9 |
| **16** | 149 | 176 | 44 | 45.8 |
| **20** | 121 | 180 | 68 | 40.2 |
| **24** | 92 | 188 | 89 | 32.9 |
| **28** | 69 | 174 | 126 | 28.4 |
| **32** | 52 | 171 | 146 | 23.3 |
| **36** | 42 | 158 | 169 | 21.0 |
| **40** | 19 | 156 | 194 | 10.9 |
| **44** | 16 | 132 | 221 | 10.8 |
| **48** | 10 | 108 | 251 | 8.5 |
| **52** | 15 | 80 | 274 | 15.8 |

***Supplemental Table 2.*** Patient primary payer type.

| **Primary payer type** | **Prescribed IpsiHand (n = 5,107)** | **Initiated therapy (n = 1,422)** | **Real-world users (n = 452)** | **With a follow-up UEFM (n = 369)** |
|---|---|---|---|---|
| Medicare | 953 (18.7%) | 705 (49.6%) | 268 (59.3%) | 208 (56.4%) |
| Medicare Advantage | 2,584 (50.6%) | 428 (30.1%) | 105 (23.2%) | 93 (25.2%) |
| Commercial / Private | 1,178 (23.1%) | 147 (10.3%) | 48 (10.6%) | 40 (10.8%) |
| Medicaid | 117 (2.3%) | 7 (0.5%) | 4 (0.9%) | 4 (1.1%) |
| Other / multiple | 1 (0.0%) | 1 (0.1%) | 1 (0.2%) | 1 (0.3%) |
| Unrecorded | 274 (5.4%) | 134 (9.4%) | 26 (5.8%) | 23 (6.2%) |
| **Medicare or Medicare Advantage (combined)** | **3,537 (69.3%)** | **1,133 (79.7%)** | **373 (82.5%)** | **301 (81.6%)** |
| **Total** | **5,107** | **1,422** | **452** | **369** |

***Supplemental Table 3.*** Breakdown of active users at each assessment window. Number of active users is defined as the number of patients with a logged BCI session within each assessment window. The denominator is the number of patients with a device during the assessment window.

| Assessment week | Real-world users, n = 452 | With a follow-up UEFM, n = 369 |
|---|---|---|
| **4** | 410/421 (97.4%) | 360/369 (97.6%) |
| **8** | 350/382 (91.6%) | 329/353 (93.2%) |
| **12** | 308/359 (85.8%) | 290/338 (85.8%) |
| **16** | 258/332 (77.7%) | 243/316 (76.9%) |
| **20** | 218/297 (73.4%) | 209/286 (73.1%) |
| **24** | 192/269 (71.4%) | 185/260 (71.2%) |
| **28** | 152/234 (65.0%) | 146/225 (64.9%) |
| **32** | 113/202 (55.9%) | 109/196 (55.6%) |
| **36** | 97/173 (56.1%) | 93/167 (55.7%) |
| **40** | 76/141 (53.9%) | 76/140 (54.3%) |
| **44** | 53/117 (45.3%) | 53/116 (45.7%) |
| **48** | 39/95 (41.1%) | 39/95 (41.1%) |
| **52** | 31/75 (41.3%) | 31/75 (41.3%) |

***Supplemental Table 4.*** Comparison of baseline characteristics between patients with and without follow-up assessments.

| | **With (Median, IQ range)** | **Without (Median, IQ range)** | **p** | **BH adjusted p** |
|---|---|---|---|---|
| **Week-12 UEFM assessment (with, n = 180; without, n = 174)** | | | | |
| Age at baseline, years | 66.0 [54.0, 72.3] | 63.5 [50.0, 71.0] | 0.17 | 0.33 |
| Baseline UEFM, 0–66 | 15.0 [8.0, 30.0] | 16.0 [10.0, 28.0] | 0.40 | 0.53 |
| Time since stroke at baseline, years | 3.5 [1.9, 6.6] | 3.5 [1.6, 6.6] | 0.54 | 0.54 |
| Social Deprivation Index, 1–100 | 41.0 [18.5, 62.5] (n = 179) | 46.0 [28.0, 68.0] | 0.072 | 0.29 |
| **Any follow-up UEFM assessment (with, n = 369; without, n = 83)** | | | | |
| Age at baseline, years | 65.0 [53.0, 72.0] | 66.0 [54.0, 72.0] | 0.60 | 0.88 |
| Baseline UEFM, 0–66 | 16.0 [9.0, 30.0] | 15.0 [9.5, 27.5] | 0.88 | 0.88 |
| Time since stroke at baseline, years | 3.4 [1.9, 6.5] | 3.9 [1.6, 7.8] (n = 76) | 0.80 | 0.88 |
| Social Deprivation Index, 1–100 | 43.5 [21.8, 65.0] (n = 368) | 37.0 [22.5, 68.0] | 0.80 | 0.88 |

#### STROBE Statement—Checklist of items that should be included in reports of cohort studies

Manuscript: “A Retrospective Real-World Assessment of Motor Status After Stroke Using a Home-Based Brain-Computer Interface.” Page and line numbers refer to the line-numbered manuscript (version of October 30, 2026); tables, figures, and supplemental items are cited by number.

| Section/Topic | Item No | Recommendation | Reported on page (lines) | Relevant text from manuscript |
|---|---|---|---|---|
| **Title and abstract** | 1 | (a) Indicate the study’s design with a commonly used term in the title or the abstract | p. 1, lines 1–4 (Title page)<br>p. 2, lines 34–37 (Abstract, Methods) | Title: “A Retrospective Real-World Assessment of Motor Status After Stroke Using a Home-Based Brain-Computer Interface”. Abstract: “This retrospective observational study included adults who were prescribed the IpsiHand System, an electroencephalography-based BCI, through a commercial US home therapy program …” |
| | | (b) Provide in the abstract an informative and balanced summary of what was done and what was found | pp. 2–3, lines 27–51 (Abstract) | Structured abstract (Background, Methods, Results, Conclusions), e.g., “At week 12, mean UEFM change was 4.3 points (95% CI, 3.7–4.9; P<0.001), and 31.7% achieved the minimal clinically important difference (MCID) of 5.25 points.” |
| **Introduction** | | | | |
| **Background/rationale** | 2 | Explain the scientific background and rationale for the investigation being reported | pp. 4–5, lines 56–100 (Introduction) | “Whether these gains are enduring, and whether benefits translate to non-clinical research settings, however, remains uncertain.” “… they were conducted using prospective protocols, and real-world evidence derived from data collected during standard clinical care remains limited.” |
| **Objectives** | 3 | State specific objectives, including any prespecified hypotheses | p. 5, lines 98–100 (Introduction)<br>p. 9, lines 190–195 (Outcome and assessment)<br>p. 6, lines 116–118 (Study design, ethics, and data governance) | “The goal of this study is to provide real-world evidence of upper extremity motor recovery with at-home BCI therapy by (1) quantifying UEFM changes after 12 weeks of use and (2) characterizing the trajectory of change through one year.” Week 12 is the prespecified primary timepoint (Outcome and assessment). |
| **Methods** | | | | |
| **Study design** | 4 | Present key elements of study design early in the paper | p. 6, lines 104–105 (Study design, ethics, and data governance, first sentence) | “This retrospective study presents a single-arm, observational characterization of the changes in UEFM within a population of real-world BCI system users.” |

| Section/Topic | Item No | Recommendation | Reported on page (lines) | Relevant text from manuscript |
|---|---|---|---|---|
| **Setting** | 5 | Describe the setting, locations, and relevant dates, including periods of recruitment, exposure, follow-up, and data collection | p. 2, lines 35–37 (Abstract, Methods)<br>p. 6, lines 109–110 (Study design, ethics, and data governance)<br>p. 6, lines 112–115 (Study design, ethics, and data governance)<br>p. 6, lines 120–122 (Patient population)<br>p. 9, lines 185–186 (Intervention, setting, and care pathway)<br>p. 9, lines 192–195 (Outcome and assessment)<br>pp. 9–10, lines 199–200 (Outcome and assessment) | Setting: a commercial US home therapy program (Abstract); "Therapy was self-administered at home, and patients chose when and how often to use the device" (Intervention, setting, and care pathway); assessments "administered remotely using videoconference, by licensed occupational therapists" (Outcome and assessment) "on a 4-week schedule" up to 52 weeks (Outcome and assessment). Dates: prescriptions from January 1, 2023, to September 6, 2026 (Patient population); "The dataset was frozen on September 6, 2026; the UEFM assessments analyzed were recorded between May 2023 and September 6, 2026" (Study design, ethics, and data governance); ethics determination dated February 7, 2025 (Study design, ethics, and data governance). |
| **Participants** | 6 | (a) Give the eligibility criteria, and the sources and methods of selection of participants. Describe methods of follow-up | p. 6, lines 120–122 (Patient population)<br>p. 7, lines 130–131 (Patient population)<br>p. 7, lines 135–136 (Patient population)<br>p. 8, lines 173–176 (Intervention, setting, and care pathway)<br>pp. 9–10, lines 198–201 (Outcome and assessment) | Inclusion: age 18 years or older and prescribed the IpsiHand within the study period, "(2) had a stroke at least six months before beginning IpsiHand therapy, and (3) had hemiparesis or hemiplegia." Exclusion: "(1) absence of a baseline UEFM measurement, (2) failure to use the device for an average of more than one session per week …", significant detrimental medical events, and cognitive, language, motor, or visual-attention impairment (Patient population). Selection through the program's care pathway and EEG screen (Patient population; Intervention, setting, and care pathway). Follow-up: "Follow-up assessments were conducted on a 4-week schedule … For analysis, follow-up assessments were organized into 4-week bins (week ± 2 weeks)." (Outcome and assessment) |
| | | (b) For matched studies, give matching criteria and number of exposed and unexposed | Not applicable | Not applicable: single-arm cohort without matching. |

| Section/Topic | Item No | Recommendation | Reported on page (lines) | Relevant text from manuscript |
|---|---|---|---|---|
| **Variables** | 7 | Clearly define all outcomes, exposures, predictors, potential confounders, and effect modifiers. Give diagnostic criteria, if applicable | p. 9, lines 189–190 (Outcome and assessment)<br>p. 10, lines 202–205 (Outcome and assessment)<br>p. 6, lines 121–122 (Patient population)<br>p. 7, lines 132–133 (Patient population)<br>p. 8, lines 152–156 (Intervention, setting, and care pathway)<br>p. 10, lines 221–223 (Statistical analysis)<br>p. 6, lines 110–112 (Study design, ethics, and data governance)<br>p. 6, lines 111–112 (Study design, ethics, and data governance) | Outcome: "The primary outcome was the UEFM total score (range 0 to 66, where higher scores indicate less impairment) during follow-up assessments up to 52 weeks"; "UEFM change was calculated as the difference between the follow-up score and the baseline score"; MCID and severity bands defined (Outcome and assessment). Exposure: IpsiHand BCI therapy (Intervention, setting, and care pathway) and device use of more than one session per week on average, counting "every logged session with a duration greater than zero" (Patient population). Covariate: "The model had two fixed effects: assessment week, entered as an unordered categorical variable, and baseline UEFM." (Statistical analysis) Characteristics: "Time since stroke was calculated from the recorded stroke date to the baseline assessment date"; SDI from ZIP code (Study design, ethics, and data governance). Diagnostic criteria: stroke at least six months earlier, hemiparesis or hemiplegia, and the definition of a significant detrimental medical event (Patient population). |
| **Data sources/measurement** | 8 | For each variable of interest, give sources of data and details of methods of assessment (measurement). Describe comparability of assessment methods if there is more than one group | p. 6, lines 107–108 (Study design, ethics, and data governance)<br>p. 9, lines 192–197 (Outcome and assessment)<br>p. 6, lines 110–112 (Study design, ethics, and data governance)<br>Supplemental Table 3 (caption) | "Data were extracted from clinical and administrative records and from device session logs." (Study design, ethics, and data governance) "Assessments were administered remotely using videoconference, by licensed occupational therapists trained and certified in the Fugl-Meyer assessment. … Training and certification included completion of the standardized BlueCloud Fugl-Meyer training, followed by at least three remotely administered Fugl-Meyer assessments reviewed and independently scored by a mentor clinician." (Outcome and assessment) "Social Deprivation Index (SDI) was determined using patient ZIP code information." (Study design, ethics, and data governance) Supplemental Table 3: active use is "a logged BCI session within each assessment window." One group, so comparability across groups does not arise. |
| **Bias** | 9 | Describe any efforts to address potential sources of bias | p. 9, lines 194–195 (Outcome and assessment)<br>p. 6, lines 116–118 (Study design, ethics, and data governance)<br>p. 11, lines 239–240 (Statistical analysis)<br>p. 12, lines 255–256 (Exploratory analyses)<br>p. 20, line 444 (Limitations) | Assessor training and certification, with assessors acknowledged as unblinded (Outcome and assessment); methods "defined prior to analysis in a statistical analysis plan" (Study design, ethics, and data governance); "Two additional sensitivity analyses were conducted to examine the missing-at-random assumption of the MMRM" (Statistical analysis); comparison of patients with and without follow-up assessments (Exploratory analyses; Supplemental Table 4); limitations (Limitations). |

| Section/Topic | Item No | Recommendation | Reported on page (lines) | Relevant text from manuscript |
|---|---|---|---|---|
| **Study size** | 10 | Explain how the study size was arrived at | p. 7, lines 133–134 (Patient population)<br>p. 13, lines 281–282 (Patient characteristics)<br>Figure 1B | “No sample size was calculated, and all eligible patients were included.” (Patient population) Of 5,107 patients prescribed IpsiHand, 1,448 received a device and were cleared to initiate therapy, and “the full patient cohort included in the analysis set consisted of 452 real-world device users who met our inclusion criteria …” (Patient characteristics; Figure 1B) |
| **Quantitative variables** | 11 | Explain how quantitative variables were handled in the analyses. If applicable, describe which groupings were chosen and why | p. 10, lines 200–201 (Outcome and assessment)<br>p. 10, lines 203–205 (Outcome and assessment)<br>p. 11, lines 233–235 (Statistical analysis)<br>pp. 6–7, lines 123–130 (Patient population)<br>p. 7, lines 130–131 (Patient population) | “For analysis, follow-up assessments were organized into 4-week bins (week ± 2 weeks).” “A ≥ 5.25 change in UEFM was chosen as the minimal clinically important difference (MCID) threshold for this study based on existing literature.” “Severity categories were determined as UEFM total score 0-28 for severe, 29-42 for moderate, and 43-66 for mild.” (Outcome and assessment) Device-use threshold: “The device-use criterion was applied so that outcomes described patients who used the device regularly.” (Patient population) Baseline UEFM enters the MMRM as a continuous covariate centered on the analysis-set mean (Statistical analysis). |
| **Statistical methods** | 12 | (a) Describe all statistical methods, including those used to control for confounding | pp. 10–12, lines 207–250 (Statistical analysis)<br>p. 12, lines 258–259 (Exploratory analyses) | Paired summaries “by the mean and its 95% confidence interval (CI) based on the t distribution, a 95% patient-level bootstrap CI”, Cohen’s dz, and paired t-tests “adjusted across all paired comparisons (n = 13) by the Benjamini-Hochberg (BH) procedure”; an MMRM with “two fixed effects: assessment week, entered as an unordered categorical variable, and baseline UEFM”, “fitted by restricted maximum likelihood”, with “Kenward-Roger degrees of freedom” (Statistical analysis); “Analyses were performed in Python 3.9.6 (pandas 2.3.3, NumPy 2.0.2, and SciPy 1.13.1) and R 4.6.1 (mmrm 0.3.18).” (Statistical analysis) Wilcoxon rank-sum tests with BH adjustment (Exploratory analyses). Confounding: no comparator group; baseline UEFM is the model covariate (Statistical analysis). |
| | | (b) Describe any methods used to examine subgroups and interactions | p. 12, lines 252–253 (Exploratory analyses)<br>p. 12, lines 260–266 (Exploratory analyses)<br>p. 20, lines 459–461 (Limitations) | “First, we compared age, baseline UEFM, time since stroke, and social deprivation index between patients that improved by at least MCID and those who did not at weeks 12, 16, and 24.” Post hoc subgroup of patients “with a baseline UEFM of 20-50, the inclusion range of previously reported clinical trials of upper extremity motor recovery after stroke” (Exploratory analyses). No interaction models: “this analysis did not model predictors of outcome” (Limitations). |
| | | (c) Explain how missing data were addressed | p. 11, lines 228–235 (Statistical analysis)<br>p. 11, lines 240–243 (Statistical analysis)<br>Table 1 | “Due to variance in follow-up trajectories, and patient factors, data were assumed missing at random and missing values were handled by direct likelihood.” Multiple-imputation sensitivity analyses (Statistical analysis). Missing characteristics shown in Table 1 as N/A rows or as the observed n. |

| Section/Topic | Item No | Recommendation | Reported on page (lines) | Relevant text from manuscript |
|---|---|---|---|---|
| | | (d) If applicable, explain how loss to follow-up was addressed | p. 11, lines 241–243 (Statistical analysis)<br>p. 12, lines 257–258 (Exploratory analyses)<br>Supplemental Table 1 | Due-but-absent assessments handled by direct likelihood under missing at random, with carry-forward multiple imputation and a tipping-point analysis (Statistical analysis); patients with and without follow-up compared (Exploratory analyses; Supplemental Table 4); Supplemental Table 1 separates observed, due-but-absent, and not-yet-due assessments at each week. |
| | | (e) Describe any sensitivity analyses | pp. 11–12, lines 239–250 (Statistical analysis) | "First, a carry-forward multiple imputation replaced each due but absent assessment with the patient's last observed value plus a zero-mean normal draw with the model's week-specific standard deviation. … Second, a tipping-point analysis … A fixed penalty term (δ) was subtracted from the imputed values after week 12 before refitting." |
| **Results** | | | | |
| **Participants** | 13* | (a) Report numbers of individuals at each stage of study—eg numbers potentially eligible, examined for eligibility, confirmed eligible, included in the study, completing follow-up, and analysed | pp. 13–14, lines 281–298 (Patient characteristics)<br>p. 14, lines 316–318 (UEFM improvements after 12 weeks)<br>Figure 1B; Supplemental Table 1 | Prescribed (5,107), received a device and cleared to initiate therapy (1,448), and analyzed: "452 real-world device users who met our inclusion criteria and received baseline UEFM assessments …, with 369 of them receiving at least one follow-up assessment" (Patient characteristics); 180 with a week-12 assessment (UEFM improvements after 12 weeks); numbers observed at each week in Supplemental Table 1. |
| | | (b) Give reasons for non-participation at each stage | p. 13, lines 282–290 (Patient characteristics)<br>Figure 1B; Supplemental Tables 1 and 3 | "Of the 3659 patients prescribed IpsiHand, but did not initiate therapy, 2225 were denied payer coverage or faced a payment barrier, 144 failed EEG screening, 695 had their inquiry case closed … and 595 were awaiting next steps …" Exclusions: 16 age, 493 no baseline UEFM, 190 chronicity, 293 device use, 4 medical events (Patient characteristics; Figure 1B). |
| | | (c) Consider use of a flow diagram | p. 14, lines 296–297 (Patient characteristics)<br>Figure 1B | Figure 1B: prescribed (5,107) → not treated (3,659, by reason) → initiated therapy (1,448) → excluded (996, by reason) → real-world users (452) → patients with follow-up UEFM (369). |
| **Descriptive data** | 14* | (a) Give characteristics of study participants (eg demographic, clinical, social) and information on exposures and potential confounders | Table 1<br>pp. 13–14, lines 291–305 (Patient characteristics)<br>Supplemental Tables 2 and 3 | Table 1: age, sex, baseline UEFM and severity band, time since stroke, affected upper extremity, dominant hand, and Social Deprivation Index, for all 452 patients and the 369 with follow-up; payer type in Supplemental Table 2; device use: "an average of 3.6 sessions per week" (Patient characteristics) and by assessment window in Supplemental Table 3. |
| | | (b) Indicate number of participants with missing data for each variable of interest | Table 1<br>p. 13, lines 293–295 (Patient characteristics)<br>Supplemental Table 4 | Table 1 lists sex (N/A, 14) and dominant hand (N/A, 65) and gives the observed n for time since stroke and the Social Deprivation Index; "253 male, 185 female of 438 with recorded sex" (Patient characteristics); Supplemental Table 4 gives the reduced n for SDI and time since stroke. |

| Section/Topic | Item No | Recommendation | Reported on page (lines) | Relevant text from manuscript |
|---|---|---|---|---|
| | | (c) Summarise follow-up time (eg, average and total amount) | pp. 13–14, lines 295–296 (Patient characteristics)<br>Supplemental Table 1 | "In these 369 patients, median follow-up from baseline to the last UEFM assessment was 16.6 weeks (IQ range [9.0, 28.1])." (Patient characteristics) Supplemental Table 1 gives, for each follow-up week, the number of patients observed, due but absent, and not yet due. |
| **Outcome data** | 15* | Report numbers of outcome events or summary measures over time | p. 15, lines 333–339 (UEFM changes over one year)<br>Supplemental Table 1; Supplemental Figure 2; Figure 3 | Observed change with SD and n at every 4-week follow-up, from "+2.4 (SD 3.6; n = 269) at week 4" to "+11.5 (SD 12.4; n = 15) at week 52" (UEFM changes over one year); model-estimated trajectory in Figure 3. |
| **Main results** | 16 | (a) Give unadjusted estimates and, if applicable, confounder-adjusted estimates and their precision (eg, 95% confidence interval). Make clear which confounders were adjusted for and why they were included | p. 14, lines 316–318 (UEFM improvements after 12 weeks)<br>p. 16, lines 348–351 (UEFM changes over one year)<br>pp. 10–11, lines 223–224 (Statistical analysis)<br>Figures 2 and 3 | Unadjusted: "a significant paired mean UEFM increase of 4.3 points (95% CI [3.7, 4.9]; bootstrap 95% CI [3.8, 5.0]; SD 4.1; …)" (UEFM improvements after 12 weeks). Adjusted for baseline UEFM, "included to account for differences in baseline impairment among the patients assessed at each week" (Statistical analysis): "Least-squares mean change from baseline was +4.2 (95% CI [3.8, 4.7]) at week 12, exceeded MCID at week 16 (+5.3; 95% CI [4.8, 5.9]), and demonstrated continued improvement up to 52 weeks (+8.1; 95% CI [6.8, 9.4])." (UEFM changes over one year) |
| | | (b) Report category boundaries when continuous variables were categorized | p. 10, lines 203–205 (Outcome and assessment)<br>p. 10, lines 200–201 (Outcome and assessment)<br>Table 1 | Severity bands 0–28, 29–42, and 43–66 (Outcome and assessment; Table 1); MCID threshold of ≥ 5.25 points (Outcome and assessment); follow-up bins of week ± 2 weeks (Outcome and assessment). |
| | | (c) If relevant, consider translating estimates of relative risk into absolute risk for a meaningful time period | Not applicable | Not applicable: continuous outcome without relative risks; proportions reaching MCID are given as absolute percentages with counts (UEFM improvements after 12 weeks). |
| **Other analyses** | 17 | Report other analyses done—eg analyses of subgroups and interactions, and sensitivity analyses | p. 15, lines 320–321 (UEFM improvements after 12 weeks)<br>p. 15, lines 328–329 (UEFM improvements after 12 weeks)<br>p. 15, lines 341–342 (UEFM changes over one year)<br>p. 16, lines 353–361 (UEFM changes over one year)<br>p. 16, lines 362–367 (UEFM changes over one year)<br>Supplemental Figures 1–6; Supplemental Tables 3 and 4 | "This pattern of continued improvement beyond week 12 persisted using the more conservative carry-forward multiple imputation approach." "The CI of the change from week 12 to week 52 included 0 at a δ of 3.0 points." (UEFM changes over one year; Supplemental Figures 5 and 6) Post hoc comparisons: Supplemental Figure 1 and Supplemental Table 4; baseline UEFM 20-50 subgroup (UEFM changes over one year). |
| **Discussion** | | | | |

| Section/Topic | Item No | Recommendation | Reported on page (lines) | Relevant text from manuscript |
|---|---|---|---|---|
| **Key results** | 18 | Summarise key results with reference to study objectives | p. 17, lines 369–375 (Discussion) | "Among patients with a paired assessment, mean UEFM improved by 4.3 points from baseline to week 12, with 31.7% of patients reaching MCID. Improvement did not plateau after 12 weeks." |
| **Limitations** | 19 | Discuss limitations of the study, taking into account sources of potential bias or imprecision. Discuss both direction and magnitude of any potential bias | p. 20, lines 444–461 (Limitations)<br>p. 16, lines 356–358 (UEFM changes over one year) | "It was observational and single-arm, without a randomized comparator." "… it does not remove the expectation effects that may accompany unblinded assessment." "Further, only patients with outcome assessments contributed to the analyses. If unassessed patients improved less, the estimates overstate improvement." (Limitations) Magnitude of departure from missing at random: "The CI of the change from week 12 to week 52 included 0 at a δ of 3.0 points." (UEFM changes over one year) |
| **Interpretation** | 20 | Give a cautious overall interpretation of results considering objectives, limitations, multiplicity of analyses, results from similar studies, and other relevant evidence | pp. 17–19, lines 376–442 (Discussion) | "Compared to previously reported UEFM outcomes in prospective and randomized clinical trials, the mean week-12 improvement in our cohort was smaller." "Although this study is observational, it establishes an initial real-world evidence base for larger, future prospective controlled trials …" Multiplicity: Benjamini-Hochberg adjustment (Statistical analysis). |
| **Generalisability** | 21 | Discuss the generalisability (external validity) of the study results | p. 20, lines 454–455 (Limitations)<br>p. 17, lines 382–384 (Discussion) | "As reflected in our cohort profile in Figure 1B, many real-world BCI users do not receive UEFM assessments, and therefore, our findings describe only patients who receive clinical follow-up services." |
| **Other information** | | | | |
| **Funding** | 22 | Give the source of funding and the role of the funders for the present study and, if applicable, for the original study on which the present article is based | p. 20, line 466 (Sources of Funding) | "This study was funded by Kandu, Inc., the manufacturer of the IpsiHand System." |

*Give information separately for exposed and unexposed groups if applicable (single-arm cohort: one group). Note: An Explanation and Elaboration article discusses each checklist item and gives methodological background and published examples of transparent reporting (von Elm et al., 2007; Vandenbroucke et al., 2007). The STROBE checklist is best used in conjunction with this article. Information on the STROBE Initiative is available at www.strobe-statement.org.
